# An optimized variant prioritization process for rare disease diagnostics: recommendations for Exomiser and Genomiser

**DOI:** 10.1101/2025.04.18.25326061

**Authors:** Isabelle B. Cooperstein, Shruti Marwaha, Alistair Ward, Shilpa N. Kobren, Jennefer N. Carter, Undiagnosed Diseases Network, Matthew T. Wheeler, Gabor T. Marth

## Abstract

**Purpose:** Whole-exome sequencing (WES) and whole-genome sequencing (WGS) are increasingly used as standard genetic tests to identify the diagnostic variants in rare disease cases. However, prioritizing these variants to reduce the time and burden of manual interpretation by clinical teams remains a significant challenge. The Exomiser/Genomiser software suite is the most widely adopted open-source software for prioritizing coding and non-coding variants. Despite its ubiquitous use, limited data-driven guidelines currently exist to optimize its performance for diagnostic variant prioritization. Based on detailed analyses of Undiagnosed Diseases Network (UDN) probands, this study presents optimized parameters and practical recommendations for deploying the Exomiser and Genomiser tools. We also highlight scenarios where diagnostic variants may be missed and propose alternative workflows to improve diagnostic success in such complex cases.

**Methods:** We analyzed 386 diagnosed probands from the UDN, including cases with coding and non-coding diagnostic variants. We systematically evaluated how tool performance was affected by key parameters, including gene:phenotype association data, variant pathogenicity predictors, phenotype term quality and quantity, and the inclusion and accuracy of family variant data.

**Results:** Parameter optimization significantly improved Exomiser’s performance over default parameters. For WGS data, the percentage of coding diagnostic variants ranked within the top ten candidates increased from 49.7% to 85.5%, and for WES, from 67.3% to 88.2%. For non-coding variants prioritized with Genomiser, the top ten rankings improved from 15.0% to 40.0%. We also explored refinement strategies for Exomiser outputs, including using *p*-value thresholds and flagging genes that are frequently ranked in the top 30 candidates but rarely associated with diagnoses.

**Conclusion:** This study provides an evidence-based framework for variant prioritization in WES and WGS data using Exomiser and Genomiser. These recommendations have been implemented in the Mosaic platform to support the ongoing analysis of undiagnosed UDN participants and provide efficient, scalable reanalysis to improve diagnostic yield. Our work also highlights the importance of tracking solved cases and diagnostic variants that can be used to benchmark bioinformatics tools.

## Background

Fewer than half of the approximately 10,000 documented rare diseases have an identified genetic etiology, however, most are presumed to be genetic in origin [1, 2]. Despite advances in next-generation sequencing, 59-75% of rare disease patients remain undiagnosed after undergoing sequencing-based tests, often due to the difficulty of accurately prioritizing and interpreting the clinical relevance of candidate variants [3–5]. Clinical diagnostic teams have limited time per patient, making it critical to minimize the number of variants requiring their review while also minimizing the chance of filtering out, and thus falsely removing from consideration, true diagnostic variants.

Variant prioritization tools integrate evidence such as population allele frequency, *in silico* predictions of variant deleteriousness, and familial segregation patterns [6]. Phenotype-based approaches refine the search further by incorporating patients’ clinical presentations encoded by Human Phenotype Ontology (HPO) [7] terms and leveraging gene:phenotype:disease associations [6]. These tools combine genotypic data in a structured framework to focus diagnostic efforts on variants most likely linked to the patient’s condition. Among freely available open-source tools for phenotype-based prioritization of single nucleotide variants (SNVs) and indels in academic and non-profit research, only Exomiser [8] (and its non-coding extension, Genomiser [9]) and AI-MARRVEL [10] met our essential selection criteria. These included accessibility via both web-based and programmatic interfaces, support for commonly used reference genomes like GRCh38, support for family-based analysis, and the ability to prioritize non-coding variants (**Supplementary Table 1**). Due to Exomiser’s more widespread use in both clinical and research settings, we selected it as the most impactful target for developing practical recommendations [5, 11, 12].

Exomiser is a scalable, secure, and efficient tool for variant prioritization, incorporating state-of-the-art annotations and prediction algorithms in regular updates. Exomiser calculates gene-level variant and phenotype scores, combining them to generate a ranked list of protein-coding candidate variants or genes across different modes of inheritance [13]. Protein-coding-focused tools such as Exomiser are the standard initial diagnostic approach due to the difficulty of interpreting variants lying outside of coding regions [14]. In contrast, the Genomiser tool was developed by the same group to focus specifically on regulatory variants, employing the same algorithms as Exomiser but expanding the search space beyond coding regions [9]. Genomiser also incorporates ReMM [9] scores, designed to predict the pathogenicity of non-coding regulatory variants. Genomiser has been shown to be effective in identifying compound heterozygous diagnoses in cases where one diagnostic variant is regulatory, and the other is coding or splice-altering [9]. However, because non-coding regions can introduce substantial noise, it has been recommended that Genomiser be used as a complementary tool alongside Exomiser rather than a replacement.

Standard user inputs to Exomiser and Genomiser include a proband or multi-sample family variant call format (VCF) [15] file, a corresponding pedigree file in PED format, and proband phenotype terms represented by HPO terms [7]. Users select tool parameters related to proband phenotype:gene similarity algorithms, variant pathogenicity scores, and frequency filters. Despite Exomiser’s widespread use, practical, data-driven guidelines for optimizing its parameters with respect to recently updated or newly developed annotations are lacking, especially for WGS data [16]. We systematically investigated the impact of these parameters on the ranking of known diagnostic variants in Exomiser and Genomiser outputs. We evaluated how differences in the quality of user-provided data, including VCF filtering criteria, HPO term quality and quantity, and familial information, impact this rank. Here, we present practical, data-supported guidelines for optimizing Exomiser and Genomiser performance using a cohort of solved cases from the Undiagnosed Diseases Network (UDN) [17]. Our study highlights scenarios where diagnostic variants may be omitted or ranked beyond the top 30 candidates and proposes alternative workflows to improve diagnostic success in complex undiagnosed cases. By applying this optimized process to an expanding number of UDN cases, we establish a scalable framework for efficient variant prioritization that can be broadly implemented for periodic reanalysis across a wide range of research and clinical scenarios.

## Methods

### UDN Participant Process and Consent

Following a comprehensive review by the clinical team, accepted UDN participants undergo indepth clinical evaluations, including WES or WGS of available affected and unaffected family members. Medical records and in-person evaluations are reviewed both manually and with computational assistance [18] to encode phenotypic features as both positive (present) and negative (absent) standardized phenotype terms using the HPO. The extensive information collected during the application and iterative evaluation process is uploaded to the UDN Data Management and Coordinating Center to support ongoing research and facilitate data sharing across the network.

UDN participants provide consent for their genomic, phenotypic, and clinical data to be shared broadly with researchers within the network. This process supports the evaluation of gene:phenotype relationships for individual probands and candidate genes and enables the use of participant data in research studies approved by the UDN Research and Publications Committee in accordance with the UDN Institutional Review Board (IRB) protocol and Manual of Operations. All UDN participant identifiers were replaced with masked IDs that do not correspond to their original consortium-assigned identifiers.

### Harmonization of exome and genome sequencing data

Multi-sample VCF [15] files for each UDN case, including the affected proband and relevant affected and unaffected family members, were extracted from cohort-level, jointly-called WGS or WES variant datasets within the UDN. These cohort-level variant datasets were produced as follows. First, unaligned, paired-end WGS FASTQ files from 5,412 unique samples corresponding to 5,353 individuals from 1,772 families enrolled in the UDN as of November 2023 were aligned to human reference GRCh38 (with decoys and all alt contigs) and processed to produce persample GVCF files in the Amazon Web Services cloud using the Clinical Genome Analysis Pipeline (CGAP, https://cgap.hms.harvard.edu/). Per-sample GVCFs were then downloaded to the local Harvard institutional cluster where SNVs and short insertions/deletions (indels) were jointly-called across all samples using Sentieon [19]. Similarly, unaligned WES FASTQ pairs from 1,454 unique samples corresponding to 1,439 individuals from 314 families enrolled in the UDN as of June 2024 were processed using the same workflow. All processing steps were performed in parallel on the Harvard institutional cluster [20].

### Curation of comprehensive phenotype lists and variant-level genetic diagnoses

HPO terms for UDN participants were stored in the UDN database by the clinical teams managing each case using PhenoTips [21]. These comprehensive HPO term lists served as input for Exomiser and Genomiser in this study. Diagnostic variants were primarily stored in HGVS format; when unavailable, they were manually extracted from sequencing reports in the UDN database. Analyses involving randomly sampled HPO terms were conducted by selecting a set of random terms from the list of all 18,697 terms in the HPO database, with each term having an equal chance of sampling, matching the proband’s comprehensive list in size but ensuring no overlaps. Analyses involving “pruned” term lists were conducted by removing all perinatal and prenatal HPO terms from probands’ comprehensive HPO term lists. A complete list of removed terms can be found in the supplementary materials.

This study considered only cases classified as having a “certain” or “highly likely” diagnosis by the UDN. Briefly, these classifications follow established diagnostic guidelines. For diseases lacking clear diagnostic criteria, diagnoses are based on a synthesis of the available objective data and the judgement of the treating clinician. A “certain” diagnosis is defined as one that is associated with minimal uncertainty, while a “highly likely” diagnosis is associated with an element of uncertainty but not enough to dismiss it for the purpose of clinical decision making [17].

### Curation of benchmarking cohorts

To establish truth sets for benchmarking Exomiser and Genomiser, we applied the following inclusion criteria to all diagnostic genes in the UDN jointly-called cohorts: (i) probands with a comprehensive list of HPO terms describing their presentation/symptoms; (ii) diagnoses labeled as “certain” or “highly likely” by the clinical team; (iii) chief method of diagnosis ‘’Genome-scale sequencing (e.g., exome sequencing, genome sequencing)” to ensure diagnosis was made primarily using sequencing data (e.g., WES or WGS) rather than other clinical tests; and (iv) SNV or indel diagnostic variants (compound heterozygous diagnoses consisting of one SNV/indel and one structural variant were excluded). Although Exomiser can handle structural variants as input, these variants were not considered as part of this study as the cohort-level harmonized variant data currently includes only SNV and indel variants. Inclusion was determined based on information retrieved from the UDN database on April 8, 2024.

Theoretically, Genomiser can be run in place of Exomiser, as it incorporates the same methods [9]. However, Genomiser’s additional consideration of non-coding variants often results in downgrading coding variants in rank. This negatively impacts its ability to prioritize coding variants, presumably due to the inherent challenges in establishing the pathogenicity of non-coding variants (**Supplementary Fig. 1**) [14]. To address this, we recommend a two-step approach when analyzing WGS variant data: using Exomiser for coding variants followed by Genomiser for non-coding variants, where necessary. Further, we curated a dedicated Genomiser benchmarking cohort focusing on WGS cases involving non-coding variants in the diagnosis.

The included data were divided into three cohorts:

1. **WES Exomiser Cohort:** WES cases with diagnostic variants in coding regions.
2. **WGS Exomiser Cohort:** WGS cases with diagnostic variants in coding regions.
3. **WGS Genomiser Cohort**: WGS cases with diagnostic variants in non-coding regions or compound heterozygous diagnoses involving non-coding variant(s).

Cohorts 1 and 2 (i.e., diagnostic coding variants) were used for Exomiser benchmarking, while cohort 3 (with diagnostic non-coding variants) was used for Genomiser benchmarking. These cohorts included probands with multiple diagnostic variants due to multiple diagnostic genes or compound heterozygous diagnoses.

Among the 181 diagnosed UDN probands with WES data, the inclusion criteria identified 153 variants in 130 genes from 125 probands (**Supplementary Fig. 2A-C**; **Table 1**). A total of 55 diagnostic genes from the WES jointly-called data were excluded for not meeting the outlined inclusion criteria or additional factors (**Supplementary Fig. 3A**). Incomplete penetrance or misphenotyping of relatives was noted in 13 families.

**Table 1:** Summary of genomic data from diagnosed UDN cases used to benchmark Exomiser and Genomiser.

| Sequencing Type | Variant Type | Probands | Diagnostic Genes | Diagnostic Variants | MOI | Test Type |
| --- | --- | --- | --- | --- | --- | --- |
| WES | Coding | 125 | 130 | 153 | AD = 90<br>AR = 54<br>XR = 2<br>XD = 7 | Singleton = 9<br>Duo = 16<br>Trio = 64<br>Quad = 26<br>>Quad = 10 |
| WGS | Coding | 231 | 239 | 296 | AD = 144<br>AR = 138<br>XR = 6<br>XD = 8 | Singleton=24<br>Duo = 29<br>Trio = 114<br>Quad = 48<br>>Quad = 16 |
| WGS | Non-coding/<br>Intronic | 39 | 39 | 60 | AD = 9<br>AR = 46<br>XR = 3<br>XD = 2 | Singleton = 1<br>Duo = 3<br>Trio = 23<br>Quad = 8<br>>Quad = 4 |

Summary of the three UDN cohorts used to benchmark Exomiser and Genomiser. MOI = Mode of Inheritance; AD = Autosomal dominant; AR = Autosomal recessive; XR = X-linked recessive; XD = X-linked dominant.

Among the 404 diagnosed UDN probands with WGS data, the inclusion criteria identified 296 variants in 239 genes from 231 probands for the WGS Exomiser Cohort. (**Supplementary Fig. 2D-F; Table 1**). The WGS Genomiser cohort included 39 probands and genes with 60 diagnostic variants (**Supplementary Fig. 2 G-I; Table 1**), including 15 genes with compound heterozygous diagnoses involving one coding and one non-coding variant. A total of 127 genes from the WGS jointly-called data were excluded for not meeting the outlined inclusion criteria or additional factors (**Supplementary Fig. 3B**). We identified inaccuracies in pedigree information, including incomplete penetrance or misphenotyping of relatives in 22 families affecting 24 diagnostic variants, leading to their exclusion from the WGS Exomiser cohort. In this context, misphenotyping refers to incorrectly annotating a family member as unaffected due to their mild phenotypic presentation.

Nine probands with coding diagnostic variants had both WES and WGS data and were evaluated in both truth sets.

### Variant filtering

Exomiser and Genomiser consider all variants present in the input VCF file as diagnostic candidates. Consequently, low-quality variants, or those likely to be artifacts of the mapping or variant calling algorithms (false positives), can be inadvertently ranked highly by these tools, increasing the burden of manual variant review. Although input VCF files can be preprocessed to remove likely false positive variants, common variant filtering strategies also risk removing true, potentially diagnostic variants from an input VCF. Standard variant callers, such as GATK [22], generate quality metrics like genotype quality (GQ), a Phred-scaled value that reflects the confidence that the genotype assigned to a sample is correct [23]. Variant allele frequency (VAF) is calculated as the proportion of sequencing reads that support an alternate allele relative to the total number of reads aligned at a specific genomic locus [23, 24]. Ideally, heterozygous loci have a VAF of 50%, homozygous alternate loci 100%, and homozygous reference loci 0%. In reality, the VAF deviates from these ideal values as a result of the sequencing process, data processing, and variant calling methods [25]. Additionally, an alternate allele value of “*” in a multi-sample VCF represents a deletion in the current sample that spans or overlaps another variant present in a different sample.

We first sought to determine optimal variant filtering criteria to minimize the inadvertent exclusion of true diagnostic variants from the input VCF. Across the three cohorts, 35 (6.9%) diagnostic variants were not prioritized using Exomiser or Genomiser default parameters using raw, unfiltered input VCFs. These variants were excluded from the filtering analysis, leaving 474 diagnostic variants to evaluate filtering criteria. Variants with an alternate allele of “*” were excluded, and the effects of filtering based on GQ and VAF of heterozygous variants were assessed. We found that increasing GQ filter stringency raised the percentage of diagnostic variants removed due to filtering (**Fig. 1A**). Narrowing the VAF window from the least (10% - 90%) to the most (25% - 75%) stringent additionally removed ∼0.5% (n=2) of diagnostic variants. As expected, the rank of diagnostic variants improved as filter stringency increased (**Fig. 1B**), highlighting the trade-off between retaining diagnostic variants and improving their prioritization rank.

**Figure 1:**
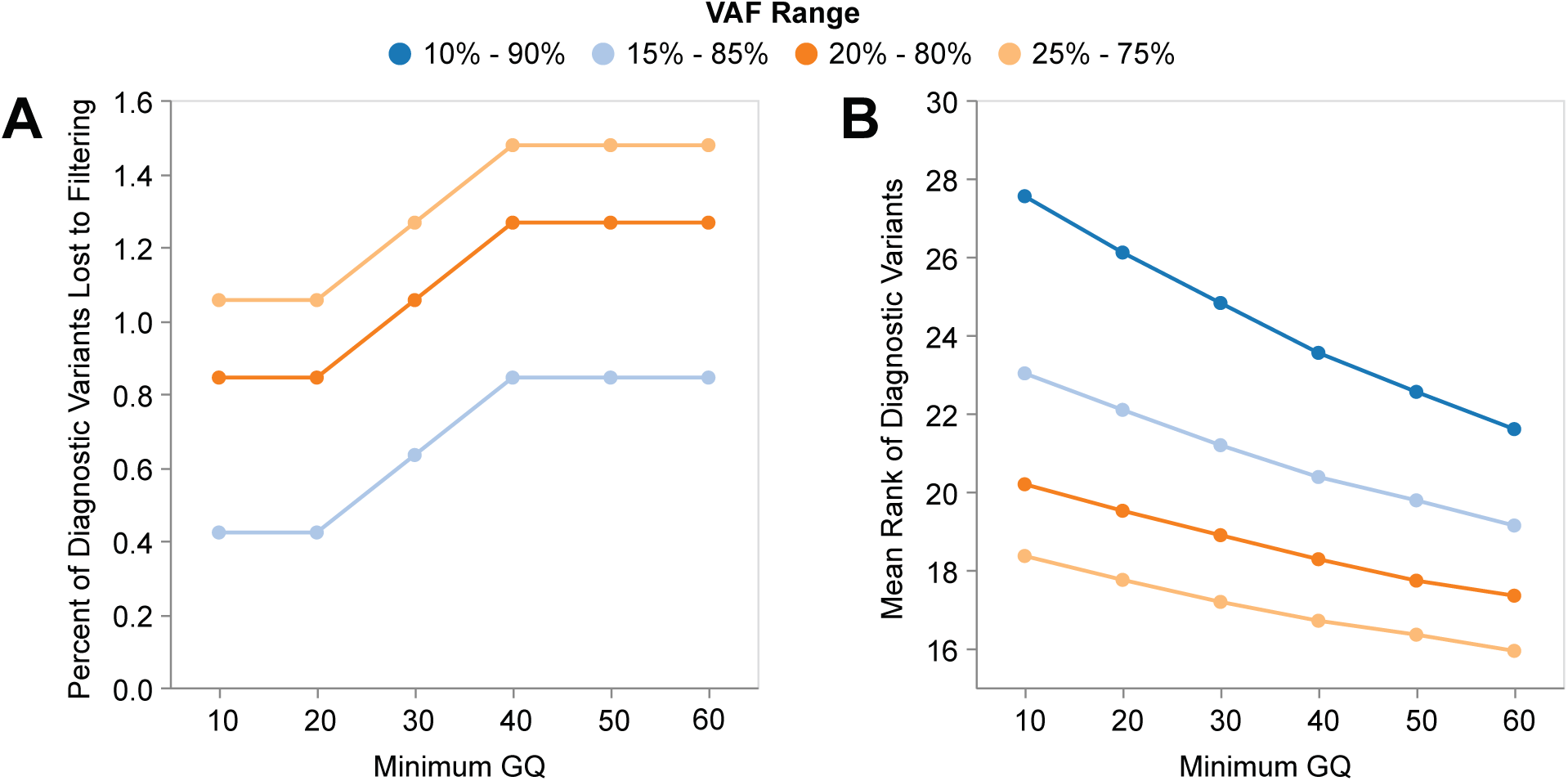
Evaluating VCF filtering criteria on 474 variants in combined WES and WGS cohorts. (A) Minimum genotype quality (GQ) versus percent of diagnostic variants removed due to filtering criteria under varying required variant allele frequency (VAF) ranges for heterozygous variants represented by colored lines. Light blue line (15%-85%) overlaps dark blue (10%-90%). (B) Minimum GQ versus mean rank of diagnostic variants in Exomiser or Genomiser outputs under default parameters with varying required VAF ranges for heterozygous variants represented by colored lines.

To balance these trade-offs, we recommend filtering input VCFs prior to variant prioritization to ensure that only high-quality variants are considered. We adopted filters of 15% ≤ VAF ≤ 85% for heterozygous variants and GQ ≥ 20 as an optimal compromise. All results in this manuscript are based on VCF files preprocessed with these filters, and all “*” alternate allele variants are removed.

We also note that filtering on VAF may lead to the exclusion of potentially diagnostic mosaic variants, which are present only in a subset of cells and, therefore, would have a VAF that deviates from 50%. This circumstance was not observed in our cohorts, as mosaic cases in the UDN were often diagnosed by methods other than standard exome or genome sequencing and were therefore excluded from this benchmarking cohort.

### Benchmarking strategy

We defined three success criteria to evaluate Exomiser and Genomiser’s ability to accurately prioritize diagnostic variants:

1. <u>Gene-level success</u>: The diagnostic gene is present in the prioritized Exomiser/Genomiser output, regardless of whether the variant position(s) or nucleotide change(s) contributing to the gene’s score are correct. This is the most lenient criterion.
2. <u>Variant-level success</u>: The diagnostic variant, including the correct position and nucleotide change, is prioritized, even if the mode of inheritance (MOI), such as autosomal dominant (AD) or autosomal recessive (AR), is incorrect. **This is our primary success measure and is most frequently reported in this manuscript.**
3. <u>Variant-level success with correct MOI</u>: The diagnostic variant meets criterion 2 and has the correct MOI, making this the most stringent criterion.

For example, in compound heterozygous cases where one variant is *not* prioritized and the second is prioritized as an AD variant, this qualifies as a gene-level success (criterion 1), one success and one failure at the variant level (criterion 2), and two failures under variant-level success with correct MOI (criterion 3).

Exomiser terminology defines the variant(s) with the highest variant score within each candidate gene as a *contributing variant*. By default, Exomiser outputs only contributing variants, even if other variants in the gene are scored. These *non-contributing* variants can be optionally included in the output. In this manuscript, a variant must be designated as a *contributing* variant to count as a variant-level success, but *non-contributing* variants are made available to users upon request via Mosaic, as described in the Results.

Finally, we imposed a top 30 ranking cutoff for success. Diagnostic variants ranked beyond 30 are unlikely to be manually reviewed and are categorized as “poor performance.”

### Bioinformatics Tools

Exomiser/Genomiser version v14.0.0 was used in the analyses reported with the dataset version 2406. Documentation and installation information can be found at https://github.com/Exomiser/Exomiser/.

All VCF subsetting, VAF calculations, and VCF filtering were done using BCFtools version 1.16 [26]. HGVS to GRCh38 genomic coordinate liftover was done using the BCFtools liftover plugin via the Broad Institute (liftover.broadinstitute.org). Exomiser/Genomiser was run using CADD v1.7 [27] and ReMM v0.4 [9] scores when applicable. The ClinVar whitelist feature was disabled for all benchmarking analyses as diagnosed UDN cases are periodically submitted to ClinVar.

### Code availability

Example YAML files and scripts used in the analyses of this paper are available in the GitHub repository at https://github.com/icooperstein/exomiser_optimization.

## Results

### Assembly of UDN benchmarking cohorts

The benchmarking cohorts comprised 386 diagnosed UDN probands and their families (<u>Methods</u>). Approximately half of the probands were diagnosed with disorders of the nervous system, including conditions of the brain and spinal cord. Probands had a median of 20 HPO terms describing their condition, with the most frequently assigned terms representing general neurological features such as *Global developmental delay* and *Seizure* (**Supplementary Fig. 3C-E**). We used these cohorts to systematically evaluate the impact of various parameters on diagnostic variant ranking by Exomiser and Genomiser, including phenotypic prioritization algorithms, variant pathogenicity prediction tools, HPO term quality and quantity, and the inclusion and accuracy of family data.

### Impact of gene:phenotype association database selection on Exomiser and Genomiser performance

Exomiser and Genomiser calculate phenotype scores using user-specified phenotypic similarity algorithms that compare proband HPO terms to known gene:phenotype associations. One option, PhenIX [28], restricts these comparisons to known Mendelian disease genes and uses gene:phenotype associations derived exclusively from human clinical data. PHIVE [29] considers all genes and uses gene:phenotype associations derived from mouse ortholog experiments. Finally, under default parameters, hiPHIVE [30] considers all genes and uses gene:phenotype associations from human, mouse, and zebrafish models, as well as phenotype associations of gene neighbors in a protein-protein interaction (PPI) network.

We compared Exomiser’s performance using PhenIX, PHIVE, default hiPHIVE, and no phenotype prioritization algorithm while keeping variant pathogenicity sources constant. As expected, omitting phenotype prioritization produced the poorest performance, with only 27.0% of diagnostic variants ranked within the top five candidates in the WGS Exomiser cohort (**Fig. 2**). Utilizing PHIVE phenotypic prioritization improved performance to 45.3%, utilizing PhenIX improved performance to 51.0%, and utilizing hiPHIVE with default parameters further improved performance to 56.8% within this range.

**Figure 2:**
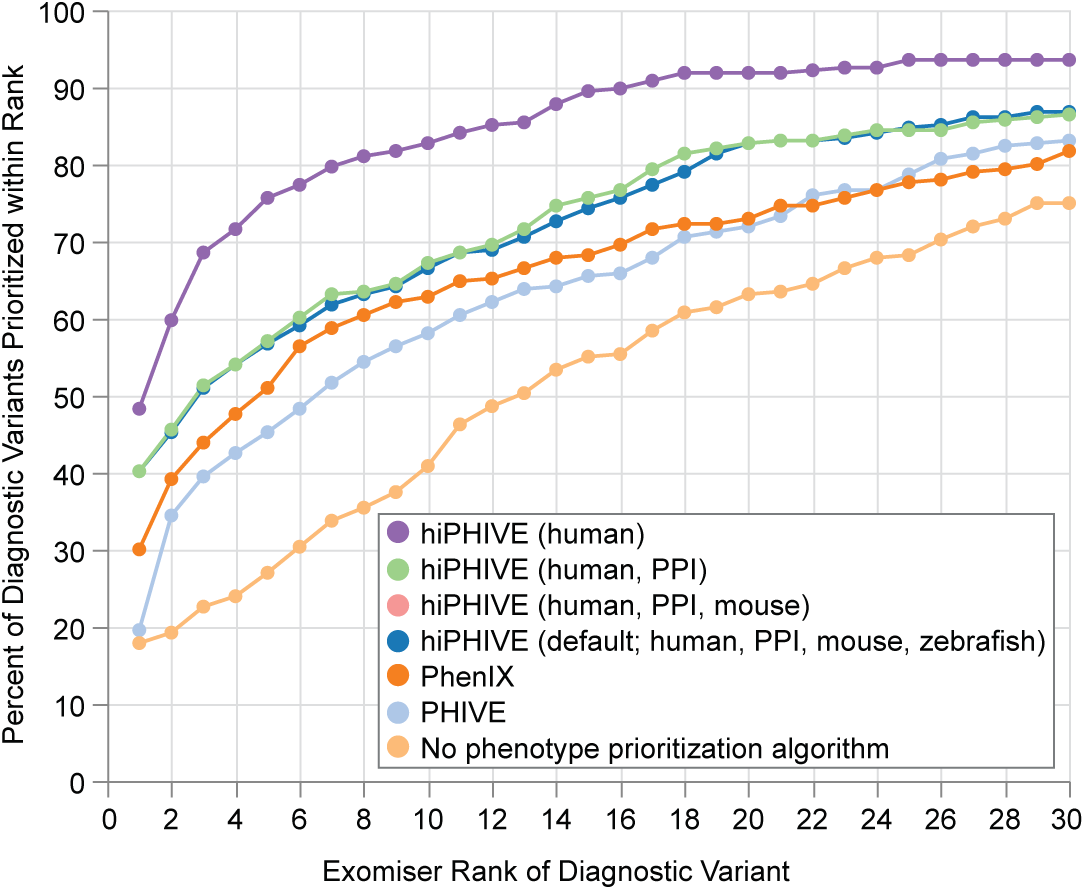
Evaluation of phenotype prioritization algorithms in the WGS Exomiser cohort. X-axis indicates the rank of the diagnostic variant in the prioritized Exomiser results. Y-axis represents the percent of diagnostic variants (out of 296) ranked at x or lower using the tested parameter type represented by color. All lines represent Exomiser run on filtered VCF using REVEL and MVP variant pathogenicity sources (default). Default phenotype prioritization algorithms (PHIVE, PhenIX, hiPHIVE) are compared, as well as the specific model organism gene:phenotype association databases used in the hiPHIVE algorithm. hiPHIVE default (dark blue) and hiPHIVE human, PPI, mouse (pink) curves overlap. X-axis is limited to 1-30 as our benchmarking strategy imposes a top 30 ranking cutoff to define success (<u>Methods</u>).

Next, we evaluated different model organism gene:phenotype association databases using the top-performing phenotype prioritization algorithm, hiPHIVE. Notably, the highest performance (75.7%) was achieved when running hiPHIVE with human-only associations (**Fig. 2**), representing over a 20% increase in performance compared to Exomiser’s recommended default hiPHIVE parameter setting using all available models of gene:phenotype associations (**Fig. 2**; **Fig. 3A; blue to purple**). Similarly, human-only hiPHIVE improved rankings in the WES cohort, with a near 15% increase in diagnostic variants within the top five candidates compared to default hiPHIVE (**Fig 3B, blue to purple**).

**Figure 3:**
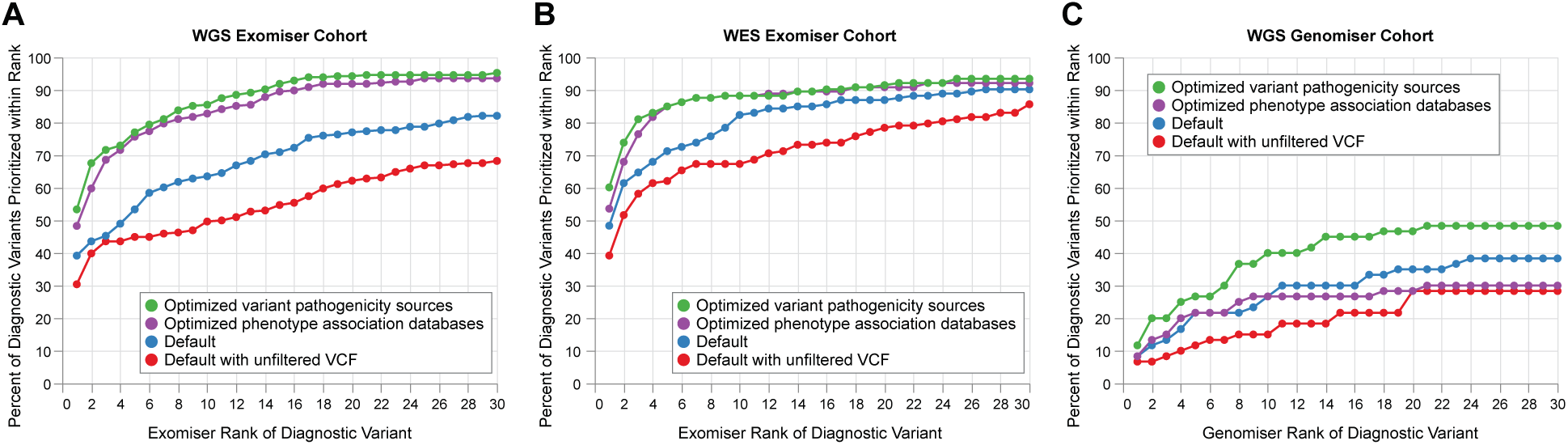
Stepwise optimization process for Exomiser and Genomiser across three UDN cohorts. (**A**) WGS Exomiser cohort (n=296 variants), (**B**) WES Exomiser cohort (n=153 variants), (**C**) WGS Genomiser cohort (n=60 variants) **Red lines**: Exomiser or Genomiser performance under default settings (hiPHIVE all models; REVEL + MVP (+ ReMM for Genomiser)) using raw, *unfiltered* VCFs. All other runs use the filtered VCFs to remove potential false positive variants (<u>Methods</u>). **Blue lines:** Exomiser or Genomiser performance under default settings (hiPHIVE all models; REVEL + MVP, (+ ReMM for Genomiser)) using filtered VCFs. **Purple lines:** Exomiser or Genomiser performance using hiPHIVE human-only associations and REVEL + MVP (+ ReMM for Genomiser) pathogenicity prediction sources. **Green lines:** Exomiser or Genomiser performance using hiPHIVE human-only associations and REVEL + MVP + AlphaMissense + SpliceAI (+ ReMM for Genomiser) pathogenicity prediction sources.

In the WGS Genomiser cohort, 21.7% of diagnostic variants were ranked within the top five candidates, regardless of whether hiPHIVE used all available models (default) or human-only gene:phenotype associations (**Fig. 3C, blue to purple**). However, for diagnostic variants prioritized under both settings, human-only hiPHIVE always yielded lower ranks (**Supplementary Fig. 4A**).

Our results indicate that restricting hiPHIVE to human-only associations improves overall ranking performance over default settings. Nevertheless, there were rare instances where non-coding diagnostic variants missed by the human-only hiPHIVE were recovered by default multispecies settings (**Supplementary Fig. 4B**). **Based on these findings, we recommend using the human-only hiPHIVE model for primary analyses due to its superior average performance. All subsequent analyses were conducted using human-only hiPHIVE.**

### Impact of variant pathogenicity prediction source selection on Exomiser performance

Each variant in Exomiser or Genomiser outputs receives a *frequency score* (0-1) based on its frequency in selected variant databases (*frequencySources*) and a *pathogenicity score* (0-1) drawn from user-specified pathogenicity predictors (*pathogenicitySources*) [16, 31]. Most of the built-in pathogenicity predictors (e.g., REVEL [32]) provide scores on a 0 (benign) to 1 (pathogenic) scale, while others (e.g., CADD [27]) are normalized by Exomiser to this range. When multiple predictors are used, Exomiser assigns the highest available score as a variant’s *pathogenicity score*. Consequently, combining many tools increases the likelihood that at least one will assign a high (pathogenic) score to a given variant, inflating scores since Exomiser uses the maximum value across sources. Variants absent from the selected pathogenicity sources are assigned a *pathogenicity score* based on variant class (**Supplementary Table 2**) [31]. The final *variant score* is the product of the *frequency* and *pathogenicity* scores. Default Exomiser parameter settings use REVEL and MVP [33] as pathogenicity sources.

We evaluated Exomiser’s average performance using individual pathogenicity sources and observed similar results across many tools (**Supplementary Fig. 5**), though differences likely exist for discrete variant classes. Adding AlphaMissense (AM) [34] and SpliceAI [35] to the default REVEL and MVP combination modestly improved performance, with AM and MVP contributing most to missense variant predictions and SpliceAI to splice variants (**Fig. 4A; Supplementary Fig. 6B**). These findings suggest that, as expected, pathogenicity prediction tools that are tailored to specific variant classes may have limited impact on overall performance but can significantly improve prioritization within the relevant subset of variants.

**Figure 4:**
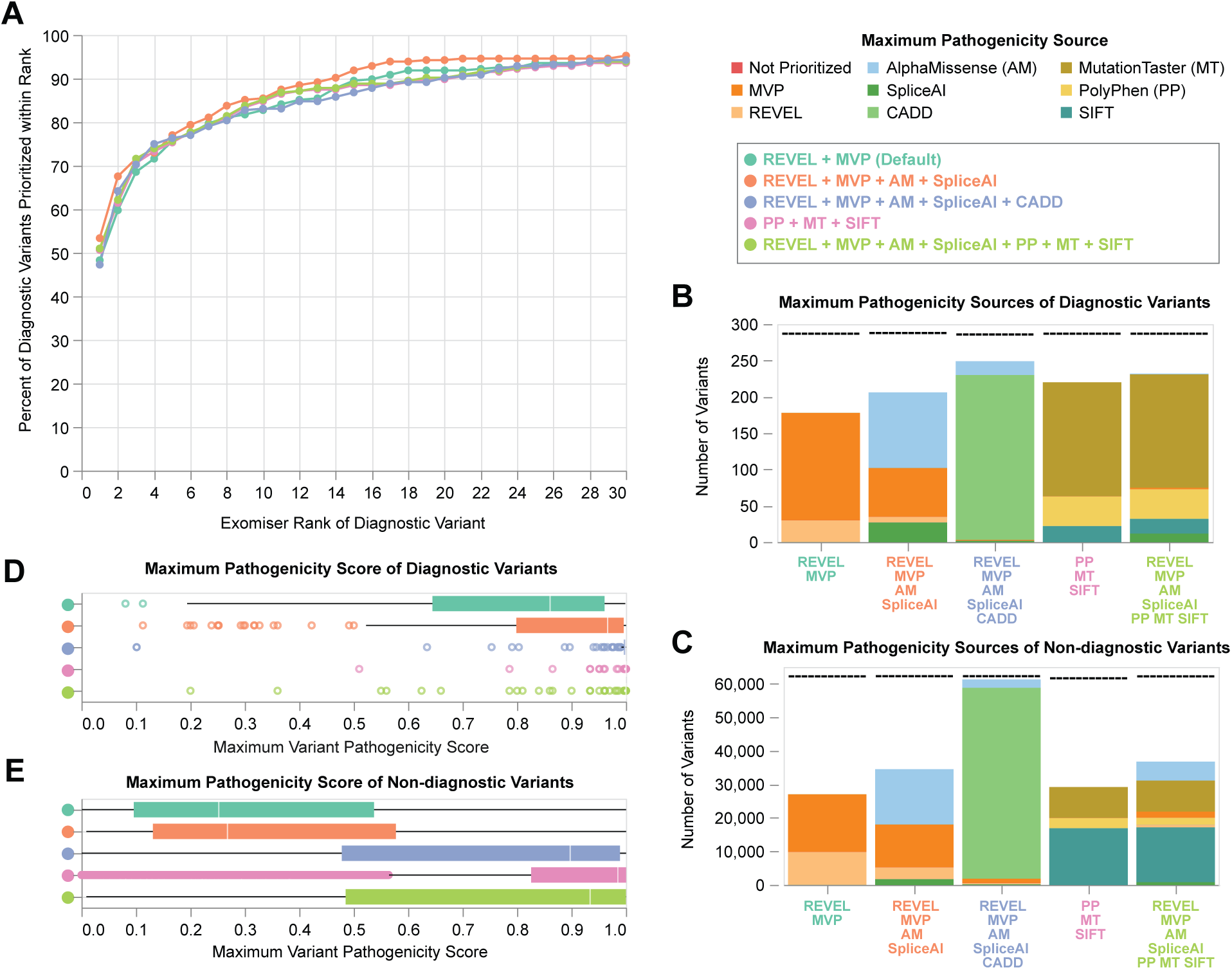
Evaluation of variant pathogenicity prediction score sources in the WGS Exomiser cohort. All data represent Exomiser run on filtered VCFs (Methods) using hiPHIVE human-only gene:phenotype annotations. (**A**) Average Exomiser performance across all variants under different combinations of variant pathogenicity prediction sources. X-axis indicates the rank of the diagnostic variant in the prioritized Exomiser results. Y-axis represents the percent of diagnostic variants (out of 296) ranked at x or lower using the selected variant pathogenicity prediction tools represented by color. Color is detailed in the lower legend. (**B-C**) Breakdown of maximum variant pathogenicity prediction score *sources* for all prioritized (**B**) diagnostic or (**C**) non-diagnostic variants under different combinations of sources (x-axis). Bar color represents the prediction source that provided the maximum pathogenicity score for the number of variants represented on the y-axis (top legend). Dashed lines represent the total number of diagnostic variants prioritized across the cohort under each run type (maximum=296 for **B**). Note that variants absent from all the selected pathogenicity sources are assigned a pathogenicity score based on their class (Supplementary Table 2) and, therefore, do not receive a maximum pathogenicity *source* (represented by this whitespace). (**D-E**) Distribution of maximum pathogenicity *scores* of all prioritized (**D**) diagnostic or (**E**) non-diagnostic variants. Color represents the combination of variant pathogenicity score sources selected in the run (lower legend).

Older tools (PolyPhen [36], MutationTaster [37], and SIFT [38]) disproportionately contributed maximum scores when included (**Supplementary Fig. 6C**), consistent with prior reports that they overestimate pathogenicity. Adding Exomiser-normalized CADD scores to REVEL, MVP, AM, and SpliceAI introduced a strong bias toward CADD as the maximum pathogenicity score source across all variant types regardless of diagnostic status (**Supplementary Fig. 6D; Fig. 4B-C**). CADD Phred-like scores reflect the log-scaled rank of a variant’s raw CADD score relative to all possible 8.6 billion SNVs in the human reference genome [27]. Exomiser’s normalization rescales these scores to a 0-1 range, effectively assigning most exonic variants - which tend to be in the top 2% of SNVs - to a score of 0.98 or higher. Because diagnostic variants are more likely to be truly pathogenic, we expect higher pathogenicity scores for these variants compared to non-diagnostic ones. This differentiation was observed using REVEL, MVP, AM, and SpliceAI but was markedly reduced when CADD or older predictors were included (**Fig. 4D-E**). Additionally, ∼10,000 variants received scores only from AM and SpliceAI, indicating broader coverage by these newer tools (**Fig. 4B-C**).

These findings suggest that normalized CADD scores bias pathogenicity rankings, likely due to difficulties in maintaining equivalency with other pathogenicity sources’ score scales, and older tools tend to overestimate pathogenicity. In contrast, including AM and SpliceAI with the default REVEL and MVP combination improves Exomiser performance, increasing the proportion of diagnostic variants ranked within the top ten by 3% over default settings (**Fig. 3A, purple to green**). **We therefore recommend the combination of REVEL, MVP, AM, and SpliceAI as pathogenicity sources for Exomiser analyses** on WGS or WES (**Supplementary Fig. 7**) variant data.

### Impact of variant pathogenicity prediction source selection on Genomiser performance

Genomiser prioritizes non-coding variants and incorporates ReMM scores, which predict the pathogenicity of non-coding regulatory variants. Genomiser’s default settings use ReMM, REVEL, and MVP as pathogenicity prediction sources. We evaluated the average performance of Genomiser across all variants using different combinations of the available sources in the WGS Genomiser cohort. Patterns were broadly consistent with those observed in the WGS Exomiser cohort, including biases introduced by older tools (PP, MT, and SIFT) and normalized CADD scores (**Supplementary Fig. 8**).

Unexpectedly, *excluding* ReMM scores improved performance in this cohort (**Supplementary Fig. 8A**). This may reflect the cohort’s small size (n=60) and lack of diagnostic variant types that ReMM is designed to predict, such as promoter or enhancer variants. Several cases involved compound heterozygous diagnoses comprising both coding (e.g., missense) and non-coding (e.g., splice-altering) variants (**Supplementary Fig. 2H**), for which ReMM is not a better predictor than missense variant prediction tools or SpliceAI. Notably, three variants (5.0%) were successfully prioritized using SpliceAI scores when ReMM was excluded but failed when ReMM was included, as higher ReMM scores were assigned to alternative variants in the diagnostic gene, resulting in the true diagnostic variants labeled as non-contributing. This suggests that for certain non-coding variant types - such as intronic splice-altering variants - tools like SpliceAI provide more informative pathogenicity predictions, but their signal can be obscured when ReMM is included due to competing high ReMM scores for other variants.

Importantly, removing ReMM is not equivalent to running Exomiser, which only considers coding variants and, in this WGS Genomiser cohort, ranked just one diagnostic variant within the top 30 candidates (**Supplementary Fig. 9**). However, this frameshift variant was incorrectly prioritized under an autosomal dominant (AD) mode of inheritance. Identifying its compound heterozygous partner —a non-coding intronic SNV— required running Genomiser. In all similar cases, Exomiser failed to prioritize the coding variant under any mode of inheritance, and Genomiser was required to recognize the pathogenicity of the coding and non-coding variant combination.

These findings highlight Genomiser’s essential role in prioritizing non-coding and regulatory variants, particularly in compound heterozygous diagnoses, which remains challenging in the field. While excluding ReMM improved performance in this specific cohort, we caution that this may reflect cohort limitations - such as the small sample size and underrepresentation of promoter or enhancer variants that ReMM is designed to predict. Given this, **we recommend running Genomiser with ReMM, REVEL, MVP, and SpliceAI**, which improved performance in the top ten candidates by 14% compared to default variant prioritization source settings (**Fig. 3C, purple to green**).

### Impact of proband phenotype term quality on Exomiser performance

HPO terms can be manually curated through clinical review or algorithmically extracted, such as via natural language processing (NLP) of medical records [18]. This variability results in some probands receiving concise terms most relevant to a genetic diagnosis, while others are assigned exhaustive lists that may include terms that are less specific or are associated with comorbidities unrelated to their genetic disease. In this study, UDN probands had a median of 20 HPO terms (range: 1-117; **Supplementary Fig. 3C**).

We evaluated how probands’ HPO term quality and quantity affected Exomiser’s ability to prioritize diagnostic variants. Removing all HPO terms significantly reduced prioritization accuracy (**Supplementary Fig. 10A**). Interestingly, assigning randomly selected HPO terms from the pool of all terms in the ontology (<u>Methods</u>) improved performance over using no terms. This likely reflects the fact that including any terms increases scores for variants in genes with known phenotypic associations, which tend to be disease-associated genes. Since most diagnosed UDN cases involve previously discovered disease-associated genes, it is unsurprising that including even randomly selected HPO terms improves performance. However, applying random terms to undiagnosed cases may bias against as-yet undiscovered disease genes and is not recommended. Additionally, randomly sampled HPO terms from the complete ontology may still be hierarchically related to probands’ phenotypes. These imprecise but nearby terms can partially capture some patients’ true phenotypic presentation, potentially inflating performance despite incorrect or overly general terms.

To assess the impact of broad term sets containing terms non-informative to the underlying genetic condition, we added randomly sampled HPO terms to probands’ comprehensive lists. Exomiser performance declined consistently with additional terms, though the effect was modest (**Supplementary Fig. 10A**). Conversely, we pruned phenotype lists for 108 probands in the WGS Exomiser cohort, removing prenatal and perinatal terms that were unlikely to be related to the underlying genetic condition (<u>Methods</u>; **Supplementary Fig. 10B**). Removing these terms changed the rank of diagnostic variants by only one position - improving rank for 17 variants (12.1%) and dropping the rank for four variants (2.9%). (**Supplementary Fig. 10D-G**).

These findings suggest that **Exomiser performs best with accurate HPO terms characteristic or indicative of genetic disorders but can tolerate imprecision that may be present in more exhaustive phenotype lists**.

### Role of pedigree accuracy and incomplete penetrance on Exomiser results

Diseases may exhibit incomplete penetrance, relatives may be misphenotyped (i.e., marked as unaffected despite mild phenotypic presentation), or pedigree files may contain human errors. We identified inaccuracies in pedigree information for 22 families with 24 diagnostic variants that led to Exomiser’s inability to rank the diagnostic variant(s) in its output. These cases were excluded from the WGS Exomiser cohort (<u>Methods</u>; **Supplementary Fig. 11A**). When reanalyzed using proband-only variant data, 21 of the 24 diagnostic variants were recovered, with eight ranked as the top candidate and 18 within the top 20 (**Supplementary Fig. 11B-C**). In many cases, manually correcting relatives’ affected status to align with the known inheritance pattern also recovered the diagnostic variants.

Notably, two diagnostic variants (8.3%) were recovered using proband-only variant data but were not prioritized when family data with corrected pedigrees were used. The true diagnoses involved single heterozygous variants consistent with AD inheritance. In both cases, Exomiser incorrectly ranked the variants under an AR model in the proband-only analysis, pairing them with additional variants in the diagnostic genes, as parental origin could not be inferred. When run with family data, the variants failed Exomiser’s inheritance filtering step as the variants did not fit a compound heterozygous model, and the variants were not recognized as damaging under an AD model.

Relatives’ phenotypic status may be uncertain or inaccurate in diagnostic analyses, hindering variant prioritization. This can be further confounded by factors like incomplete penetrance and multigenic disorders with overlapping phenotypes. **These findings highlight the importance of accurate pedigree information and suggest that re-evaluating the affected status of family members may help recover diagnostic variants within the top 30 ranked candidates**. We also assessed how family variant data and the inheritance filtering step influence Exomiser’s ability to prioritize diagnostic variants when pedigree information is accurate. This analysis confirmed that leveraging family data improves performance but also highlighted the inheritance filtering step as essential for enhancing prioritization even when only proband information is available (**Supplementary Material**; **Supplementary Fig. 12**).

### Strategies to refine candidate variant lists from Exomiser and Genomiser outputs

A major goal of variant prioritization is to reduce the number of candidate variants requiring manual review while retaining the diagnostic variant. In this study, we optimized Exomiser and Genomiser parameters to improve the ranking of diagnostic variants, ideally within the top 30 candidates. However, some diagnostic variants may still fall above this threshold and require additional support from other ‘omic data, such as RNA or long-read sequencing. Ideally, this multiomic data should be analyzed together. In such cases, a broader list of candidate variants may be helpful. For example, this list may be cross-referenced against genes that show expression or splicing outliers, or structural variants in the same gene that may suggest compound heterozygosity.

Although Exomiser provides *p*-values for each ranked variant and gene, there are no established guidelines for using these values to filter candidate lists. We explored whether *p*-value thresholds could serve as an alternative to a fixed top 30 cutoff to generate a less stringent prioritized list of SNV/indel variants for downstream integration with other data types. We evaluated multiple thresholds (**Supplementary Fig. 13**) and found that *p* ≤ 0.3 offered a favorable balance - reducing noise, decreasing the median number of prioritized variants per proband, and excluding only 5 (1.7%) diagnostic variants in the WGS Exomiser cohort (**Supplementary Table 3**). However, the number of retained variants was strongly influenced by family structure, with singleton cases yielding more candidates under the same threshold as compared to trios. While the *p*-value cutoff may reduce noise in integrated analyses, we caution that the ideal threshold for multi-omics applications likely depends on the specific context and analysis framework. Additional benchmarking is needed to define best practices for integrating Exomiser results with other ‘omic data.

Additionally, we observed that certain genes regularly contributed high-scoring but non-diagnostic variants across this cohort of patients with very diverse phenotypes. To reduce noise and ensure a focus on high-quality candidates, we first applied a *p*-value threshold of *p* ≤ 0.3. Across 231 WGS Exomiser cases, 86 genes appeared in the top 30 with *p* ≤ 0.3 in at least 5% of probands (**Fig. 5**). Removing variants in these genes from the Exomiser output and subsequent reranking resulted in the loss of 14 (4.7%) diagnostic variants but moderately improved the rank of 78 (26.4%) (**Supplementary Fig. 14B-C**). We also identified frequently ranked genes in the WES Exomiser cohort (**Supplementary Fig. 15**), but the small sample size of the WGS Genomiser cohort (n=39 probands) precluded a similar analysis, as 5% of this benchmarking cohort represents only two cases. Removing variants from genes that are frequently prioritized among the top candidates by Exomiser in a given cohort may result in the loss of true diagnostic variants. Therefore, **we suggest a tiered approach that flags genes with frequently “over-prioritized” variants that can be further manually interpreted and eliminated cautiously.**

**Figure 5:**
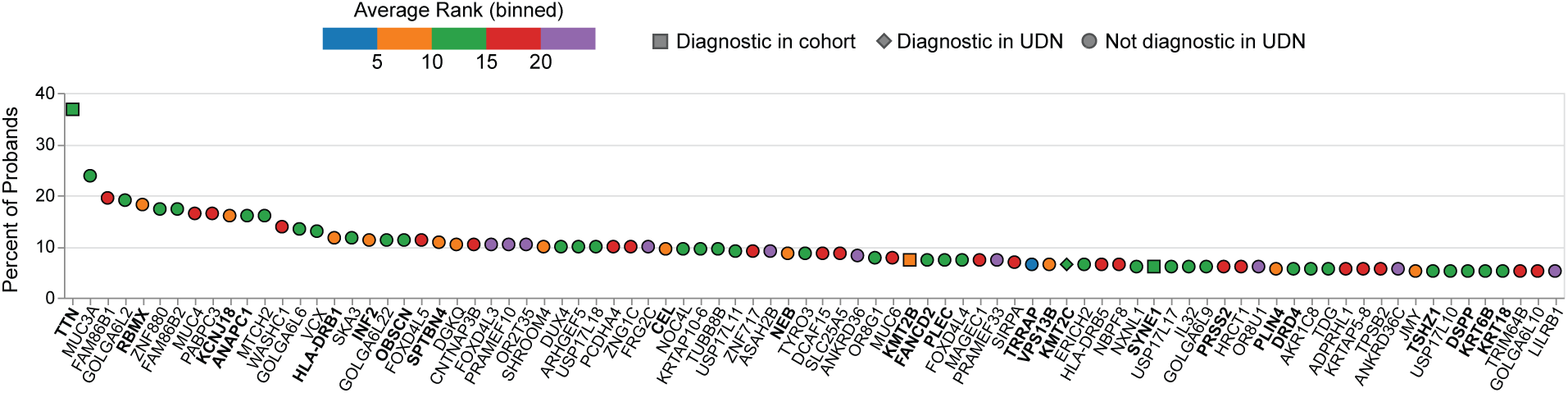
Frequently ranked genes in the WGS Exomiser cohort. 86 genes that ranked in the top 30 candidates with *p* ≤ 0.3 for at least 5% of probands in the WGS Exomiser cohort. Color represents the binned average rank at which the gene was prioritized across all probands in the cohort. Bold font indicates OMIM genes with a confirmed causal relationship to a disease (OMIM 3). Shape reflects if the gene is diagnostic in the WGS Exomiser cohort (square), not diagnostic in the WGS Exomiser cohort but diagnostic for at least one proband in the UDN consortia as a whole (diamond), or not diagnostic in the UDN consortia (circle).

### Applying optimized Exomiser parameters for variant reanalysis

Exomiser’s fast runtime and strong performance in ranking diagnostic variants using optimized parameters make it well-suited for variant reanalysis in undiagnosed patients, particularly in response to evolving clinical phenotypes. We therefore integrated our Exomiser guidelines into the ongoing analysis of undiagnosed UDN cases via the Mosaic platform.

Mosaic is a genomic data management and analysis platform used by UDN researchers that supports collaborative diagnostics via user-friendly interfaces. The user interface allows users to search approximately 200,000 to 400,000 variants per UDN family. All Exomiser rankings and scores generated via the best practices outlined in this paper are made available. When participants undergo a change in phenotype, updated Exomiser scores are made available based on the updated HPO term list.

## Discussion

Exomiser and Genomiser are complementary open-source tools designed to prioritize coding and non-coding variants in genomic data, respectively. In this study, we used diagnosed cases from the Undiagnosed Diseases Network (UDN) to evaluate the performance of these tools, propose best practices, and identify potential pitfalls in rare disease diagnostics. Optimizing Exomiser parameters significantly improved its performance over default settings, with 85.5% of diagnostic variants in the WGS Exomiser cohort ranked in the top ten candidates compared to 63.5% under default settings after applying our recommended filtering criteria (<u>Methods</u>; **Fig. 3A**). This optimization shifted 70 (23.6%) variants into the top ten candidates (**Fig. 6**), representing a substantial improvement for manual review by elevating variants that may have otherwise been missed at higher ranks. Across all diagnostic variants in this cohort, optimization improved the rank for 152 variants and did not change the rank for 125. Although the rank of 19 diagnostic variants increased, none were displaced from the top 30 candidates. This reflects an inherent tradeoff, where improvements in overall performance may occasionally deprioritize certain variants. Similar improvements were observed for WES data and non-coding variants prioritized using Genomiser (**Fig. 3B-C; Supplementary Fig. 16A-B**).

**Figure 6:**
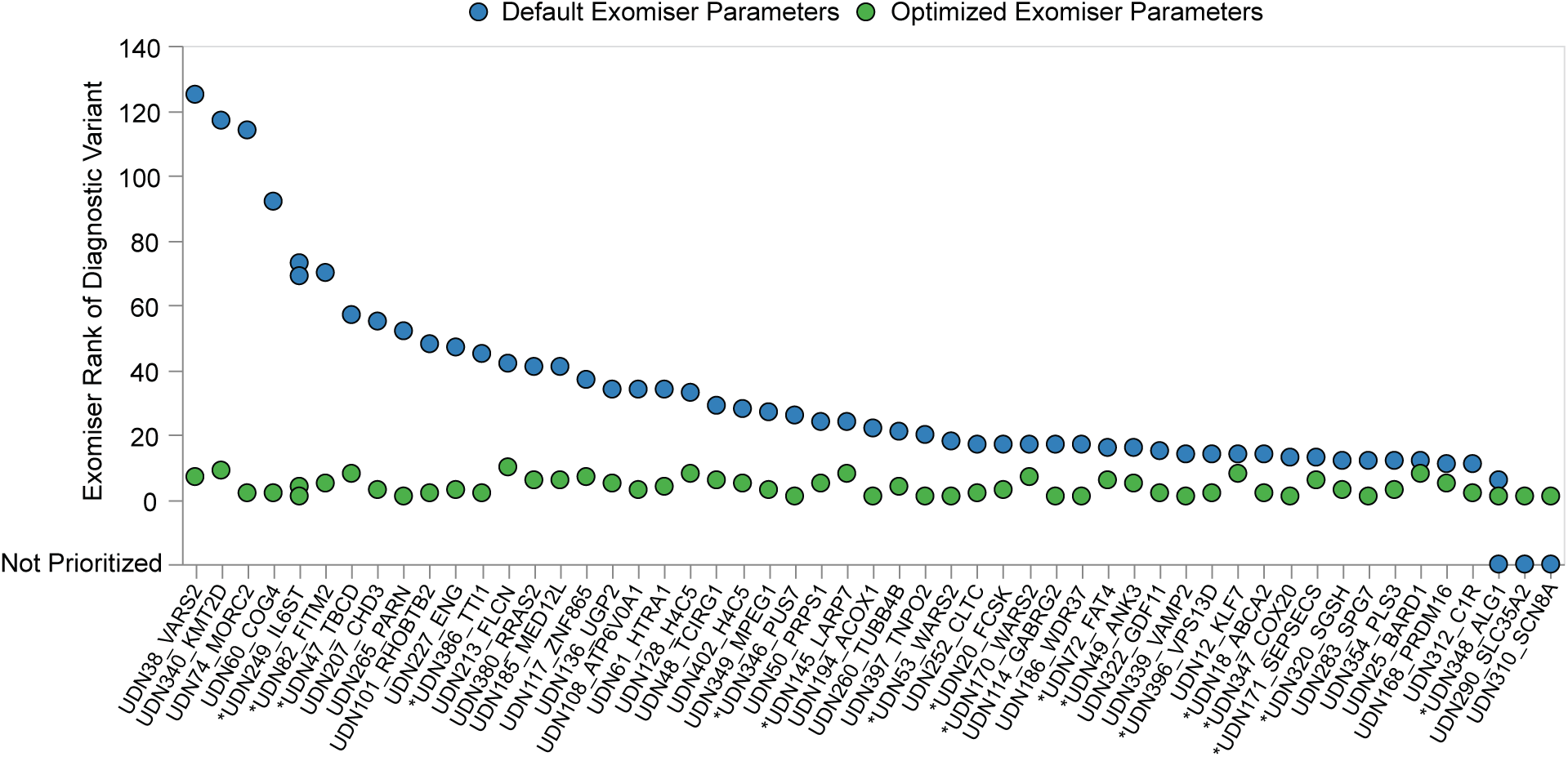
Parameter optimization shifts diagnostic variants into the top ten candidates in the WGS Exomiser cohort. 70 (23.6%) variants in the WGS Exomiser cohort are shifted into the top ten candidates using optimized parameters (green) in comparison to default parameters (blue). Optimized parameters refer to running Exomiser on the *filtered* family VCF, hiPHIVE human-only gene:phenotype associations and REVEL, MVP, AlphaMissense, and SpliceAI variant pathogenicity score sources. Default parameters refer to running Exomiser on the filtered family VCF using hiPHIVE human, mouse, zebrafish, and PPI gene:phenotype associations and REVEL and MVP variant pathogenicity score sources. * denotes compound heterozygous diagnosis (2 variants in labeled genes).

Based on the analyses performed in this study, we recommend the following best practices for using Exomiser/Genomiser in rare disease diagnostics (**Fig. 7**):

1. Begin analysis with a family (jointly-called) VCF (when available) and filter it to remove potential false positive variants (<u>Methods</u>)
2. Use the REVEL, MVP, AlphaMissense, and SpliceAI variant pathogenicity prediction sources and human-only hiPHIVE gene:phenotype associations
3. Utilize pedigree information, Exomiser inheritance filters, and enable the ClinVar whitelist*
4. Manually review the top 30 *contributing* variants

**Figure 7:**
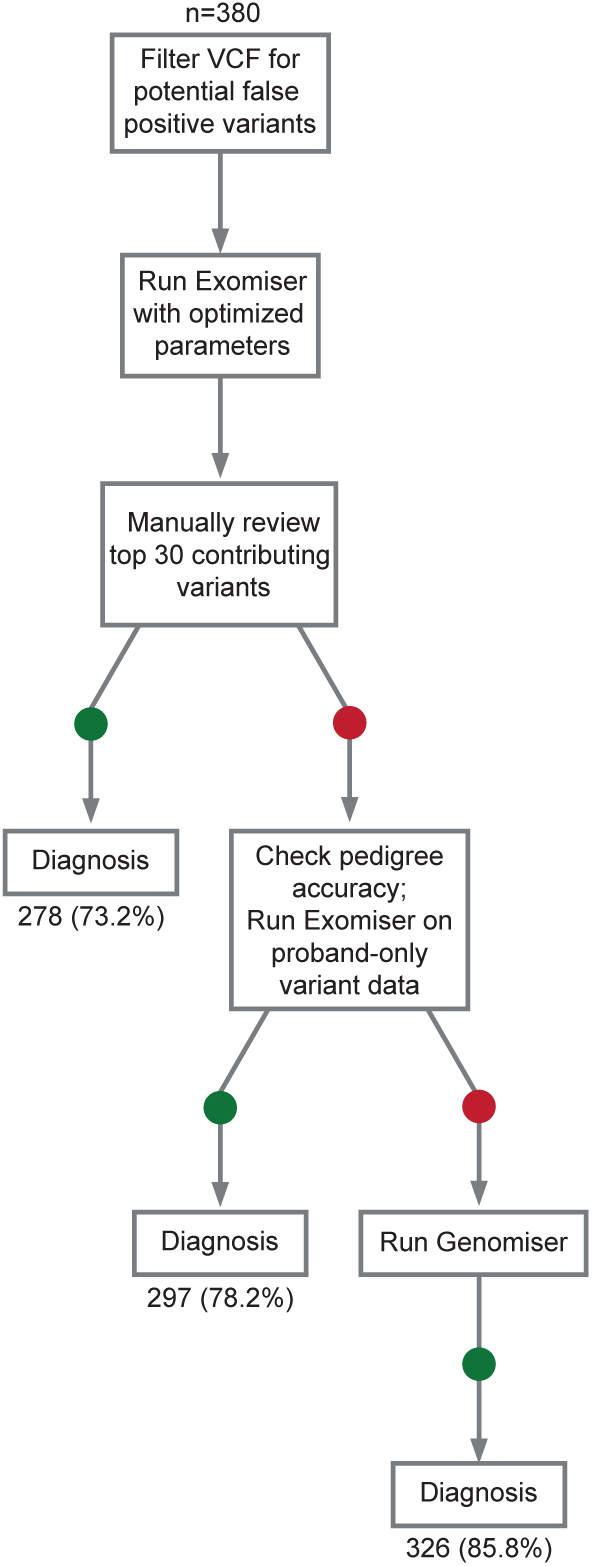
Recommended workflow for using Exomiser and Genomiser in rare disease diagnostics. Numbers indicate the count (percentage) of diagnostic variants ranked *within the top 30 candidates* by Exomiser or Genomiser after applying each preceding step in the flowchart. Percentages are based on n=380 diagnostic variants (296 from the WGS Exomiser cohort, 24 with inconsistent pedigrees, and 60 from the WGS Genomiser cohort). Begin analysis with a family VCF (when available), filtered to remove potentially false positive variants (<u>Methods</u>). Use the REVEL, MVP, AlphaMissense, and SpliceAI variant pathogenicity sources and human-only hiPHIVE gene:phenotype associations. Run Exomiser using all available family variant data, pedigree information, inheritance filters, and the ClinVar whitelist enabled. Manually review the top 30 *contributing* variants, with frequently prioritized genes flagged. If no compelling candidates are identified, verify pedigree accuracy pedigree information (considering that some family members may be misphenotyped) and consider running Exomiser on the proband-only variant data with inheritance filters enabled. If no strong candidates are found in WGS data, run Genomiser to assess non-coding variants and compound heterozygous candidates with one non-coding variant and one coding variant. Additional optional analyses include running Exomiser and/or Genomiser with multispecies and PPI gene:phenotype associations and applying *p*-value thresholds to generate broader gene lists for integration with other ‘omic datasets.

*Any variant on the ClinVar whitelist bypasses all Exomiser filters (e.g., allele frequency) and will be included in the prioritized results with a maximal score. Although we excluded this feature to avoid biasing benchmarking results, we observed improved performance when enabled, so we recommend its inclusion when analyzing unsolved cases. We also found that Exomiser performs best with accurate and relevant HPO terms but tolerates some imprecision or noise in phenotype lists.

There are scenarios in which the outlined steps do not prioritize the diagnostic variant(s), prompting us to introduce additional analyses that can be explored to further improve outcomes when time permits (**Fig. 7**). First, we observed that some of the diagnostic variants were missed by Exomiser due to inheritance constraints resulting from pedigree inaccuracies (**Supplementary Fig. 11**). We also demonstrated that Genomiser, while requiring more manual curation, is necessary for identifying non-coding candidates. In cases with compound heterozygous diagnoses made up of non-coding and coding variants, Exomiser alone was unable to detect both diagnostic variants in the correct AR configuration (**Supplementary Fig. 9**). However, while Genomiser considers coding variants, it is not a replacement for Exomiser, as it identified ∼30% fewer diagnostic variants within the top 30 candidates and required nearly three times the runtime (**Supplementary Fig. 1**).

While restricting gene:phenotype associations to human-only data improved overall performance, in rare instances, it also led to missed variants in genes with recently established disease associations not yet represented in human-specific databases. Expanding to include multispecies and PPI data enabled recovery of such variants, as seen for six variants in the WGS Genomiser cohort (**Supplementary Fig. 4**). In a separate subset, Exomiser failed to prioritize diagnostic variants within the top 30 candidates (**Supplementary Table 4**). These instances of poor performance, based on our criteria for success, were typically driven by low phenotype or variant scores. Together, these findings underscore the importance of regular updates to and the expansion of gene:phenotype association databases and variant pathogenicity predictors to improve prioritization outcomes, particularly in challenging or novel cases.

We encountered a specific challenge with using CADD scores in Exomiser and Genomiser. To be incorporated, CADD Phred scores are normalized to a 0-1 range, where a score of 10 maps to 0.9. However, in our experience, a REVEL score of 0.9 indicates pathogenicity more than a CADD Phred score of 10, particularly for missense variants. As Exomiser assigns the highest score across variant pathogenicity sources, these normalized CADD scores disproportionately bias maximum pathogenicity scores due to their incompatibility with other sources’ scoring scales. As a result, we excluded CADD scores from our analyses. This decision does not reflect a flaw in CADD or Exomiser but rather highlights a limitation in the compatibility of these scales. We recommend that users review CADD scores alongside Exomiser results, as they provide valuable context for variant interpretation. Future updates to Exomiser that improve variant pathogenicity scale equivalency may mitigate this issue.

Finally, we explored refinement strategies for Exomiser outputs, including filtering by *p*-value and identifying genes frequently ranked in the top 30 candidates but rarely associated with diagnoses. These approaches can support hypothesis generation in complex or unsolved cases by enabling integration of prioritized SNV/indels with other variant types (e.g., structural variants) or ‘omes (e.g., expression or splicing outliers), or by flagging variants that analysts should interpret with caution, informed by trends observed across a larger benchmarking cohort.

We acknowledge some limitations in this study. The cohort size, particularly for Genomiser benchmarking, was relatively small, which may limit the generalizability of our findings. Additionally, we recognize the biases inherent in solved UDN cases, where well-documented gene:phenotype associations may be more available and could have led to more favorable outcomes compared to unsolved cases. Furthermore, some variant pathogenicity prediction tools, such as REVEL, are trained on ClinVar data which may include diagnostic variants in this cohort, potentially inflating their ability to predict these variants as damaging.

## Conclusions

This study provides valuable insights into the capabilities and limitations of Exomiser and Genomiser for variant prioritization in rare disease diagnostics. By optimizing parameters and refining analysis strategies, we significantly improved tool performance in prioritizing diagnostic variants as compared to default settings. Our findings underscore the importance of integrating informative patient phenotypes and accurate family pedigrees with high-quality variant data. This work also suggests evidence-based best practices for deploying these tools for rare disease diagnostics. While the optimized parameters enhance diagnostic outcomes, there are cases where they may fall short, highlighting the continued necessity of manual review in clinical applications. Our work also highlights the significance of tracking and documenting diagnosed variants that can be used for benchmarking bioinformatic pipelines.

## Data Availability

All deidentified exomic and genomic sequencing data including SNV and indel variant calls, as well as corresponding phenotype data in the form of pedigree files and HPO terms, are regularly deposited in dbGaP (accession phs001232.v5.p2). Rare SNV and indel variants and HPO terms for UDN participants with genomic sequencing are also queryable in a public-facing browser: https://dbmi-bgm.github.io/udn-browser/. Variant-level data, clinical significance and supporting evidence, demographic information, and phenotype information for all diagnostic variants, including those included in the WES Exomiser, WGS Exomiser, and WGS Genomiser datasets used in this study, are submitted to ClinVar. Other relevant, patient-specific clinical information may be shared on a case-by-case basis at the discretion of the clinical team managing the case.

https://github.com/icooperstein/exomiser_optimization

## List of abbreviations

WES: Whole-exome sequencing
WGS: Whole-genome sequencing
UDN: Undiagnosed Diseases Network
HPO: Human Phenotype Ontology
SNV: Single nucleotide variant
VCF: Variant call format
IRB: Institutional Review Board
GQ: Genotype quality
VAF: Variant allele frequency
MOI: Mode of inheritance
AD: Autosomal dominant
AR: Autosomal recessive
PPI: Protein-protein interaction
AM: AlphaMissense
NLP: Natural language processing

## Declarations

### Ethics approval and consent to participate

All work included in this study was performed in accordance with all ethical guidelines outlined in the NIH IRB #15HG0130 and the UDN Manual of Operations. All deidentified patient data included in this study was provided with informed consent by all participants to be used freely for research purposes across the network. The study proposal and this manuscript were approved by the UDN Publications and Research Committee.

## Consent for publication

Not applicable.

## Authors’ contributions

SM, IC, and AW contributed towards conception and design of the study including methods and interpretation of the results. SM piloted benchmarking of Exomiser and Genomiser using a small subset of solved cases from the UDN. IC scaled analysis to all diagnosed cases for results in this manuscript. IC and SM fetched metadata associated with participants. IC curated benchmarking cohorts. SK performed realignment and joint calling of UDN WGS and WES data. IC performed parameter optimizations, computational analyses, and completed all figure generation. AW ran Exomiser and Genomiser on unsolved UDN cases and facilitated their availability in Mosaic. IC, SM, and AW drafted the original manuscript. SK, JC, MW and GTM contributed to manuscript review and editing. All authors read and approved the final manuscript.

## Acknowledgements

We thank the Exomiser development team for making Exomiser and Genomiser freely available and for their ongoing work to support and improve rare disease diagnostics through open-source tools.

## References

1. Haendel M, Vasilevsky N, Unni D, et al (2020) How many rare diseases are there? Nat Rev Drug Discov 19:77–78

2. Ferreira CR (2019) The burden of rare diseases. American Journal of Medical Genetics Part A 179:885–892

3. Chung CCY, Hue SPY, Ng NYT, Doong PHL, Hong Kong Genome Project, Chu ATW, Chung BHY (2023) Meta-analysis of the diagnostic and clinical utility of exome and genome sequencing in pediatric and adult patients with rare diseases across diverse populations. Genet Med 25:100896

4. Wright CF, Campbell P, Eberhardt RY, et al (2023) Genomic Diagnosis of Rare Pediatric Disease in the United Kingdom and Ireland. N Engl J Med 388:1559–1571

5. 100,000 Genomes Project Pilot Investigators, Smedley D, Smith KR, et al (2021) 100,000 Genomes Pilot on Rare-Disease Diagnosis in Health Care - Preliminary Report. N Engl J Med 385:1868–1880

6. Jacobsen JOB, Kelly C, Cipriani V, Research Consortium GE, Mungall CJ, Reese J, Danis D, Robinson PN, Smedley D (2022) Phenotype-driven approaches to enhance variant prioritization and diagnosis of rare disease. Hum Mutat 43:1071–1081

7. Robinson PN, Mundlos S (2010) The Human Phenotype Ontology. Clin Genet 77:525–534

8. Smedley D, Jacobsen JOB, Jäger M, et al (2015) Next-generation diagnostics and disease-gene discovery with the Exomiser. Nat Protoc 10:2004–2015

9. Smedley D, Schubach M, Jacobsen JOB, et al (2016) A Whole-Genome Analysis Framework for Effective Identification of Pathogenic Regulatory Variants in Mendelian Disease. Am J Hum Genet 99:595–606

10. Mao D, Liu C, Wang L, et al (2024) AI-MARRVEL - A Knowledge-Driven AI System for Diagnosing Mendelian Disorders. NEJM AI. 10.1056/aioa2300009

11. Laurie S, Piscia D, Matalonga L, et al (2022) The RD-Connect Genome-Phenome Analysis Platform: Accelerating diagnosis, research, and gene discovery for rare diseases. Hum Mutat 43:717–733

12. Buske OJ, Girdea M, Dumitriu S, et al (2015) PhenomeCentral: a portal for phenotypic and genotypic matchmaking of patients with rare genetic diseases. Hum Mutat 36:931–940

13. Smedley D, Robinson PN (2015) Phenotype-driven strategies for exome prioritization of human Mendelian disease genes. Genome Med 7:81

14. Belkadi A, Bolze A, Itan Y, Cobat A, Vincent QB, Antipenko A, Shang L, Boisson B, Casanova J-L, Abel L (2015) Whole-genome sequencing is more powerful than whole-exome sequencing for detecting exome variants. Proc Natl Acad Sci U S A 112:5473–5478

15. Danecek P, Auton A, Abecasis G, et al (2011) The variant call format and VCFtools. Bioinformatics 27:2156–2158

16. Cipriani V, Pontikos N, Arno G, et al (2020) An Improved Phenotype-Driven Tool for Rare Mendelian Variant Prioritization: Benchmarking Exomiser on Real Patient Whole-Exome Data. Genes (Basel). 10.3390/genes11040460

17. Splinter K, Adams DR, Bacino CA, et al (2018) Effect of Genetic Diagnosis on Patients with Previously Undiagnosed Disease. N Engl J Med 379:2131–2139

18. Deisseroth CA, Birgmeier J, Bodle EE, et al (2019) ClinPhen extracts and prioritizes patient phenotypes directly from medical records to expedite genetic disease diagnosis. Genet Med 21:1585–1593

19. Freed D, Aldana R, Weber JA, Edwards JS (2017) The Sentieon Genomics Tools - A fast and accurate solution to variant calling from next-generation sequence data. bioRxiv. 10.1101/115717

20. Kobren SN, Moldovan MA, Reimers R, et al (2024) Joint, multifaceted genomic analysis enables diagnosis of diverse, ultra-rare monogenic presentations. bioRxiv 2024.02.13.580158

21. Girdea M, Dumitriu S, Fiume M, et al (2013) PhenoTips: patient phenotyping software for clinical and research use. Hum Mutat 34:1057–1065

22. Van der Auwera GA, O’Connor BD (2020) Genomics in the Cloud: Using Docker, GATK, and WDL in Terra. O’Reilly Media

23. Carson AR, Smith EN, Matsui H, Brækkan SK, Jepsen K, Hansen J-B, Frazer KA (2014) Effective filtering strategies to improve data quality from population-based whole exome sequencing studies. BMC Bioinformatics 15:125

24. Boscolo Bielo L, Trapani D, Repetto M, Crimini E, Valenza C, Belli C, Criscitiello C, Marra A, Subbiah V, Curigliano G (2023) Variant allele frequency: a decision-making tool in precision oncology? Trends Cancer Res 9:1058–1068

25. Kraft IL, Godley LA (2020) Identifying potential germline variants from sequencing hematopoietic malignancies. Hematology Am Soc Hematol Educ Program 2020:219–227

26. Danecek P, Bonfield JK, Liddle J, et al (2021) Twelve years of SAMtools and BCFtools. Gigascience. 10.1093/gigascience/giab008

27. Schubach M, Maass T, Nazaretyan L, Röner S, Kircher M (2024) CADD v1.7: using protein language models, regulatory CNNs and other nucleotide-level scores to improve genome-wide variant predictions. Nucleic Acids Res 52:D1143–D1154

28. Zemojtel T, Köhler S, Mackenroth L, et al (2014) Effective diagnosis of genetic disease by computational phenotype analysis of the disease-associated genome. Sci Transl Med 6:252ra123

29. Smedley D, Oellrich A, Köhler S, Ruef B, Sanger Mouse Genetics Project, Westerfield M, Robinson P, Lewis S, Mungall C (2013) PhenoDigm: analyzing curated annotations to associate animal models with human diseases. Database 2013:bat025

30. Bone WP, Washington NL, Buske OJ, et al (2016) Computational evaluation of exome sequence data using human and model organism phenotypes improves diagnostic efficiency. Genet Med 18:608–617

31. Robinson PN, Köhler S, Oellrich A, et al (2014) Improved exome prioritization of disease genes through cross-species phenotype comparison. Genome Res 24:340–348

32. Ioannidis NM, Rothstein JH, Pejaver V, et al (2016) REVEL: An Ensemble Method for Predicting the Pathogenicity of Rare Missense Variants. Am J Hum Genet 99:877–885

33. Qi H, Zhang H, Zhao Y, Chen C, Long JJ, Chung WK, Guan Y, Shen Y (2021) MVP predicts the pathogenicity of missense variants by deep learning. Nat Commun 12:510

34. Cheng J, Novati G, Pan J, et al (2023) Accurate proteome-wide missense variant effect prediction with AlphaMissense. Science 381:eadg7492

35. Jaganathan K, Kyriazopoulou Panagiotopoulou S, McRae JF, et al (2019) Predicting Splicing from Primary Sequence with Deep Learning. Cell 176:535–548.e24

36. Adzhubei IA, Schmidt S, Peshkin L, Ramensky VE, Gerasimova A, Bork P, Kondrashov AS, Sunyaev SR (2010) A method and server for predicting damaging missense mutations. Nat Methods 7:248–249

37. Schwarz JM, Cooper DN, Schuelke M, Seelow D (2014) MutationTaster2: mutation prediction for the deep-sequencing age. Nat Methods 11:361–362

38. Ng PC, Henikoff S (2001) Predicting deleterious amino acid substitutions. Genome Res 11:863–874

39. Arriaga MT, Mendez R, Ungar RA, et al (2025) Transcriptome-wide outlier approach identifies individuals with minor spliceopathies. medRxiv. 10.1101/2025.01.02.24318941

