## Supplementary Materials for "An optimized variant prioritization process for rare disease diagnostics: recommendations for Exomiser and Genomiser"

#### Impact of the inclusion of family variant data and inheritance filtering step on Exomiser performance

Exomiser's inheritance filtering step uses information about the affected status of available family members as well as variant zygosity, making it valuable even when only proband data is available. Applying this step using all available family variant data yields the most accurate results (**Supplementary Fig. 12**). However, the inheritance filtering step significantly enhances prioritization accuracy whether family variant data is available or not. Proband-only variant data with inheritance filtering outperforms family variant data without this step, as Exomiser considers the family data only when the inheritance filtering step is applied.

Without inheritance filtering, Exomiser cannot detect autosomal recessive (AR) patterns. Fifty-four AR variants were prioritized using proband-only variant data with the inheritance filtering step applied but were missed when this step was skipped. Including family variant data and applying this filtering step enabled prioritization of two additional AR variants.
These findings confirm that ***leveraging family data improves Exomiser's performance, but also highlight the inheritance filtering step as essential for fully leveraging family information and enhancing prioritization when only proband data is available***. However, this inheritance-aware analysis is limited to the variants and variant types represented in the input VCF. As shown by the need for Genomiser to detect compound heterozygous diagnoses involving coding and non-coding variants, similar limitations exist for cases involving a combination of one SNV/indel and one structural variant (SV). We excluded such cases from this study, as SVs were not represented in the available VCFs. While recent versions of Exomiser support SV prioritization, future benchmarking efforts will be needed to evaluate its performance in this context.

#### Diagnostic variants not prioritized by Exomiser in the WGS Exomiser cohort

Eight variants (2.7%) in the WGS Exomiser cohort failed Exomiser prioritization as defined by our *variant-level success* criterion, as described in Methods (**Supplementary Table 5**). It is important to note that these are edge cases and not limitations of Exomiser. However, the user should be aware of such scenarios.
Two of these variants (<1%) were filtered out of the input VCF due to high VAF values. These variants were called with a 1/2 genotype, indicating that the proband inherited two different alternate alleles from each parent at that position. However, a standard bioinformatic pipeline step is the decomposition of variants, which split this variant into two records in the VCF with incorrect 1.0 VAF values. This resulted in both variants being filtered out by our imposed $0.15 \leq VAF \leq 0.85$ filtering step. Without this filtering step, these diagnostic variants remain in the VCF and correctly rank as the top candidate.
Five variants (1.7%) failed at least one Exomiser-imposed filtering step. Under our recommended parameters, the Exomiser frequency filter step only passes variants with a minor allele frequency (MAF) < 2% across all frequency databases. Three diagnostic variants failed this step. The other two variants were filtered out by Exomiser's inheritance filtering step: one

due to exceeding the maximum MAF threshold for AD inheritance (0.1%), and the other due to failure to recognize a compound heterozygous AR configuration with a second variant in the same gene.
The final variant which failed at the variant-level success criterion passed all filtering steps but was reported as non-contributing. Although the correct diagnostic gene was ranked as the top candidate, the true variant's pathogenicity score was slightly lower than that of an alternative AD candidate in the same gene. This underscores the importance of retaining non-diagnostic variants in output files, as they may gain diagnostic relevance under alternative analyses.

#### Perinatal/prenatal HPO terms removed to create "pruned" term lists

HP:0012188 | Hyperemesis gravidarum
HP:0008071 | Maternal hypertension
HP:0009800 | Maternal diabetes
HP:0030244 | Maternal fever in pregnancy
HP:0100622 | Maternal seizure
HP:0011438 | Maternal teratogenic exposure
HP:0100603 | Toxemia of pregnancy
HP:0011436 | Abnormal maternal serum screening
HP:0001511 | Intrauterine growth retardation
HP:0001562 | Oligohydramnios
HP:0001561 | Polyhydramnios
HP:0001558 | Decreased fetal movement
HP:0010519 | Increased fetal movement
HP:0001787 | Abnormal delivery
HP:0001622 | Premature birth
HP:0001518 | Small for gestational age
HP:0001520 | Large for gestational age
HP:0003561 | Birth length less than 3rd percentile
HP:0003517 | Birth length greater than 97th percentile
HP:0011451 | Primary microcephaly
HP:0004488 | Macrocephaly at birth
HP:0002643 | Neonatal respiratory distress
HP:0006579 | Prolonged neonatal jaundice
HP:0002033 | Poor suck
HP:0001998 | Neonatal hypoglycemia
HP:0040187 | Neonatal sepsis
HP:0011410 | Caesarian section
HP:0030364 | Secondary Caesarian section
HP:0001787 | Abnormal delivery
HP:0030369 | Induced vaginal delivery
HP:0001788 | Premature rupture of membranes
HP:0100603 | Toxemia of pregnancy
HP:0001194 | Abnormalities of placenta or umbilical cord

### Supplementary Figures

A

B

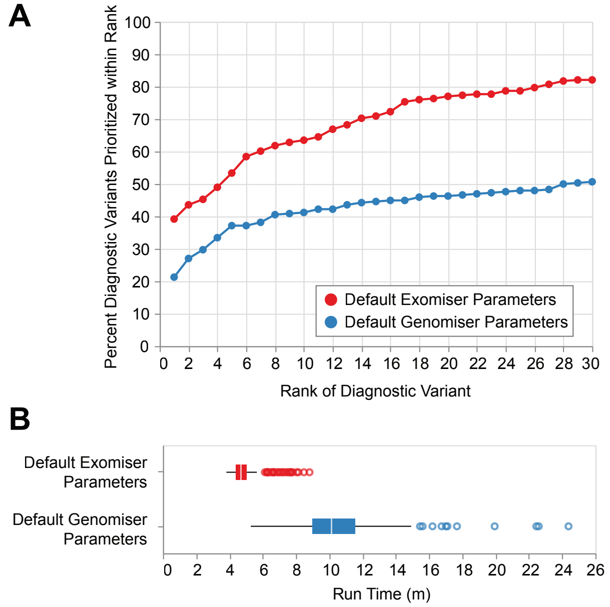


**Supplementary Figure 1: Comparison of Exomiser and Genomiser performance on coding variants.**

For completeness, we evaluated the performance of Genomiser on prioritizing **coding** variants relative to Exomiser. Genomiser identified approximately 30% fewer diagnostic variants within the top 30 candidates and required nearly three times the run time in comparison to Exomiser on the same cohort.

(**A**) Default Exomiser and Genomiser performance on filtered VCFs from the WGS Exomiser (n=296 variants).

(**B**) Run time in minutes for Exomiser and Genomiser using default parameters on filtered VCFs from the WGS Exomiser cohort (n=231 probands).

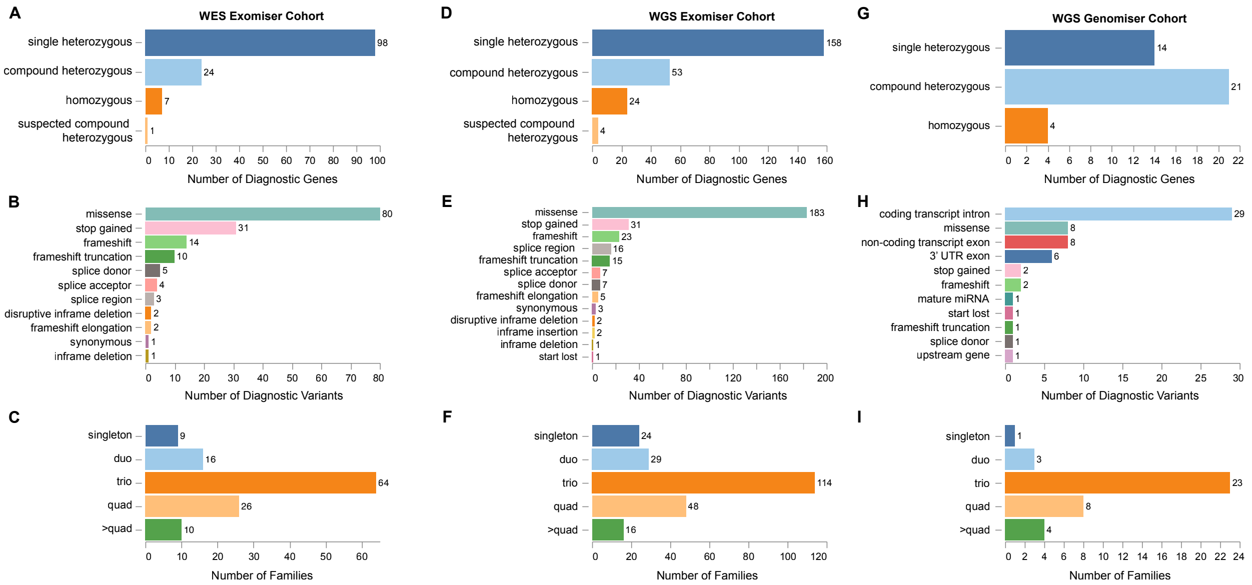


**Supplementary Figure 2: Breakdown of three UDN cohorts used in benchmarking.**
**Left panel:** WES Exomiser cohort. Breakdown of (**A**) variant zygosity in 130 diagnostic genes, (**B**) 153 diagnostic variant types, (**C**) family structures of 125 probands in jointly-called WES variant data that met inclusion criteria.
**Middle panel**: WGS Exomiser cohort. Breakdown of (**D**) variant zygosity in 239 diagnostic genes, (**E**) 296 diagnostic variant types, (**F**) family structures of 231 probands in jointly-called WGS variant data that met inclusion criteria.
**Right panel:** WGS Genomiser cohort. Breakdown of (**G**) variant zygosity in 39 diagnostic genes, (**H**) 60 diagnostic variant types, (**I**) family structures of 39 probands in jointly-called WGS variant data that met inclusion criteria.

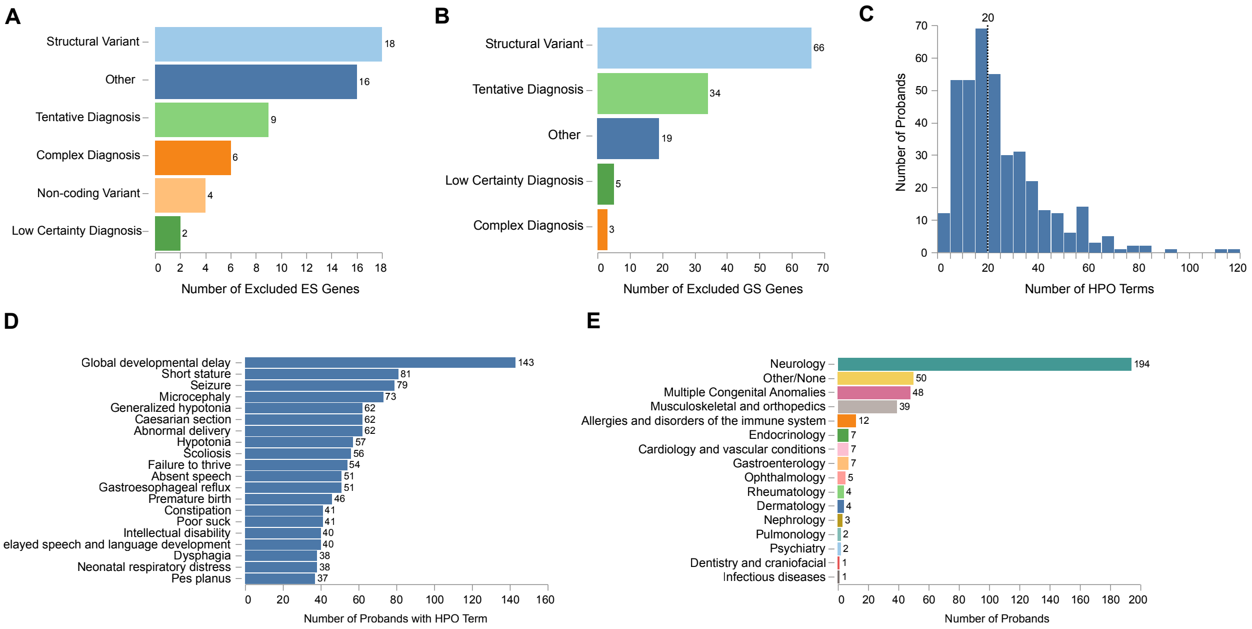


**Supplementary Figure 3: Summary of combined WES/WGS cohorts.**

Combining all WES and WGS data resulted in 386 unique probands (125 WES Exomiser + 231 WGS Exomiser + 39 WGS Genomiser – 9 probands with both WES and WGS)

(**A**) Criteria for genes excluded from the WES cohort analyses (n=55).

(**B**) Criteria for genes excluded from the WGS cohort analyses (n=127).

“Other” in **A/B** includes complex diagnoses (multiple gene interactions), mitochondrial variants, the absence of diagnostic variant(s) in provided VCF, non-sequencing primary diagnosis methods, and one case where no second variant was ever found so diagnosis was considered incomplete.

(**C**) Distribution of the number of HPO terms in probands’ comprehensive HPO list from the UDN database. Dashed line represents the median number of terms per proband in 386 probands in combined WES/WGS cohorts.

(**D**) Twenty most commonly assigned HPO terms in combined WES/WGS cohorts.

(**E**) Distribution of primary symptom categories assigned to probands in combined WES/WGS cohorts.

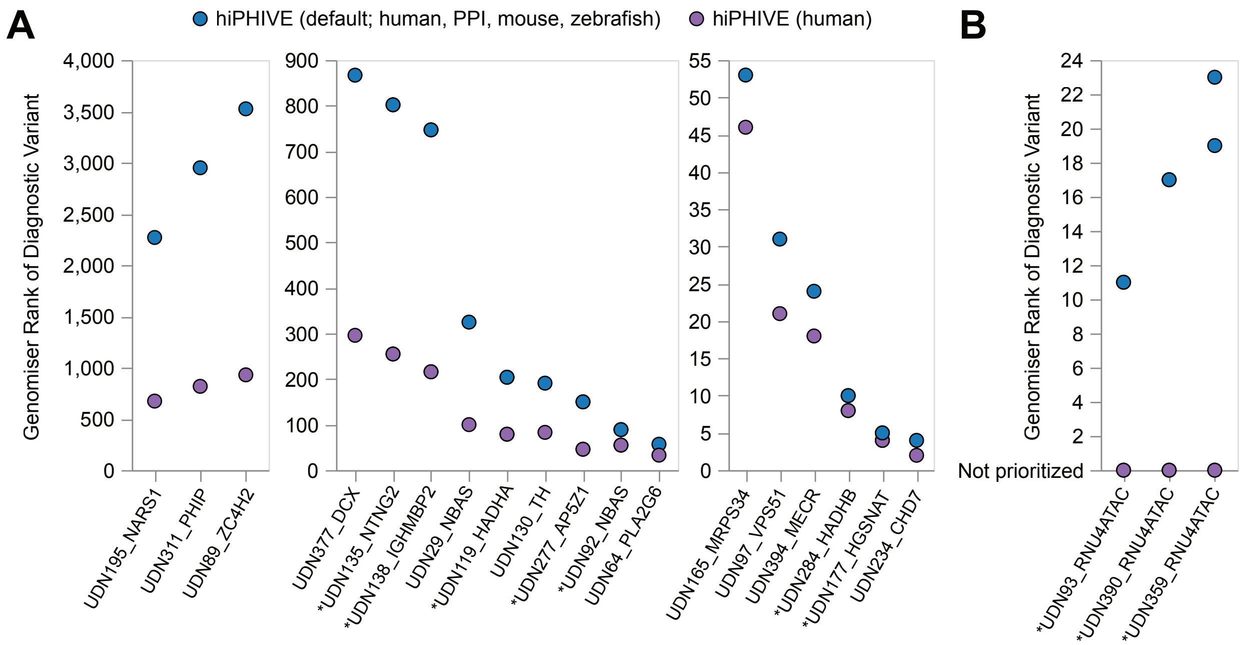


**Supplementary Figure 4: Genomiser performance using hiPHIVE with default versus human-only gene:phenotype associations.**

Performance of Genomiser was tested using gene-specific phenotype scores calculated using either the hiPHIVE algorithm with all models (default; human, zebrafish, mouse, PPI databases) of gene:phenotype associations (blue) or human-only associations (purple).

(**A**) 25 (41.7%) variants changed rank when run using all models of gene:phenotype associations (blue) versus human-only gene-phenotype associations (purple). Variants always showed improved rank when Genomiser was run using human-only gene:phenotype associations (purple).

Notably, the default hiPHIVE's average performance (**Fig. 3C, blue**) appears inflated due to six variants that fall in the small non-coding RNA gene RNU4ATAC, which has only recently been found to be highly associated in humans with neurodevelopmental disorders [39]. These variants failed prioritization with human-only hiPHIVE but were prioritized under default hiPHIVE.

(**B**) 6 (10%) variants that were *not* prioritized when run using human-only gene:phenotype associations (purple) but were prioritized when using all models of gene-phenotype associations (blue).

* denotes compound heterozygous diagnosis (2 variants in labeled genes).

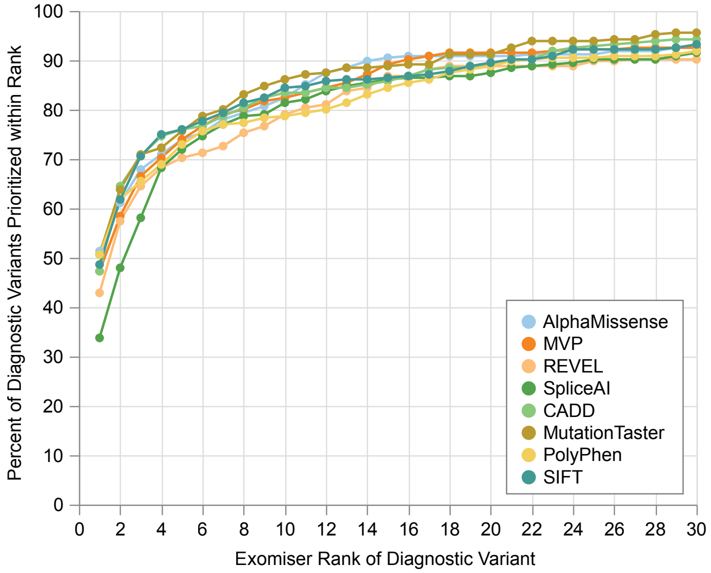


**Supplementary Figure 5: Evaluation of individual variant pathogenicity prediction score sources in WGS Exomiser cohort.**

The performance of individual variant pathogenicity prediction sources available to run Exomiser with was tested. X-axis indicates the rank of the diagnostic variant in the prioritized Exomiser results. Y-axis represents the percent of diagnostic variants (out of 296) that are ranked at x or lower using the selected variant pathogenicity prediction tool represented by color. All lines represent Exomiser run on filtered VCFs (Methods) using hiPHIVE human-only gene:phenotype annotations. X-axis is limited to 1-30 as our benchmarking strategy imposes a top-30 ranking cutoff to define success (Methods).

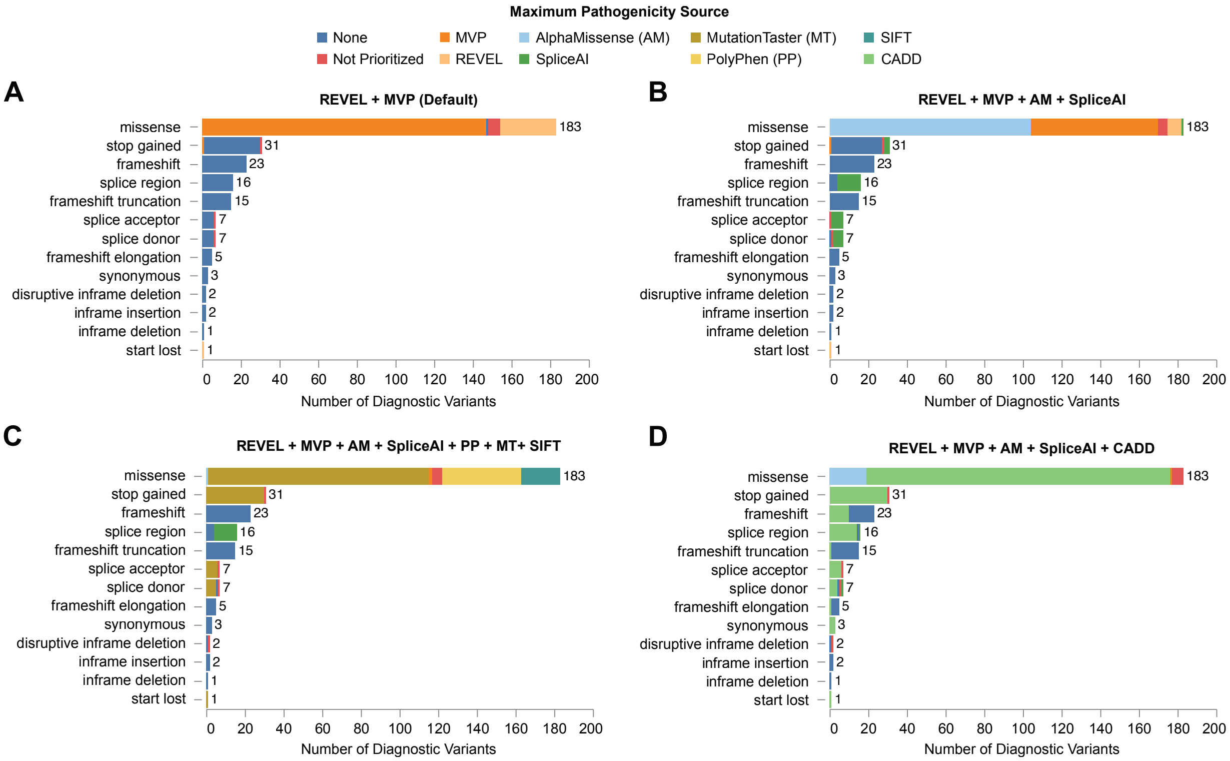


**Supplementary Figure 6: WGS Exomiser diagnostic variants' maximum pathogenicity score source broken down by variant class.**

X-axes represent the number of variants for each variant class on the y-axis (totaling to 296). Bar color represents the prediction source that provided the maximum pathogenicity score for each variant under different source combinations listed in title. "None" represents variants that were prioritized, but did not receive a score from any of the selected pathogenicity score sources.

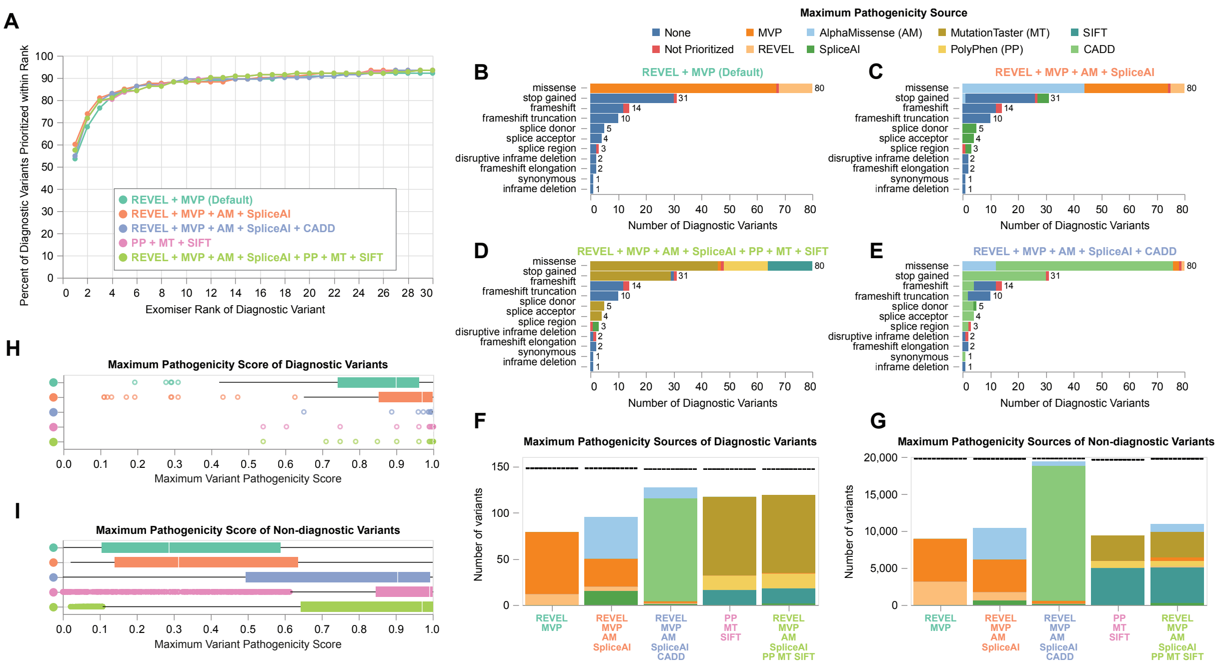


**Supplementary Figure 7: Evaluation of variant pathogenicity prediction score sources in WES Exomiser cohort.**

All data represent Exomiser run on filtered VCFs using hiPHIVE human-only gene:phenotype annotations.

(**A**) Average Exomiser performance across all variants under different combinations of variant pathogenicity prediction sources. X-axis indicates the rank of the diagnostic variant in the prioritized Exomiser results. Y-axis represents the percent of diagnostic variants (out of 153) that are ranked at x or lower using the selected variant pathogenicity prediction tools represented by color.

(**B-E**) Diagnostic variants' maximum pathogenicity score source broken down by variant class. X-axes represent the number of variants for each variant class on the y-axis (totaling to 153). Bar color represents the prediction source that provided the maximum pathogenicity score for each variant under different source combinations listed in title (top right legend). "None" represents variants that were prioritized, but do not receive a score from any selected pathogenicity score sources.

(**F-G**) Breakdown of maximum variant pathogenicity prediction score *sources* for all prioritized (**F**) diagnostic or (**G**) non-diagnostic variants under different combinations of sources (x-axis, legend in **A**). Bar color represents the prediction source that provided the maximum pathogenicity score for the number of variants represented on the y-axis (top right legend). Dashed lines represent the total number of diagnostic variants prioritized across the cohort under each run type (maximum=153 for **F**). Note that variants absent from all the selected pathogenicity sources are assigned a pathogenicity score based on their class (Supplementary Table 2), and therefore do not receive a maximum pathogenicity *source* (represented by this whitespace).

(**H-I**) Distribution of maximum pathogenicity *scores* of all prioritized (**H**) diagnostic or (**I**) non-diagnostic variants. Color represents the combination of variant pathogenicity score sources selected in run (legend in **A**).

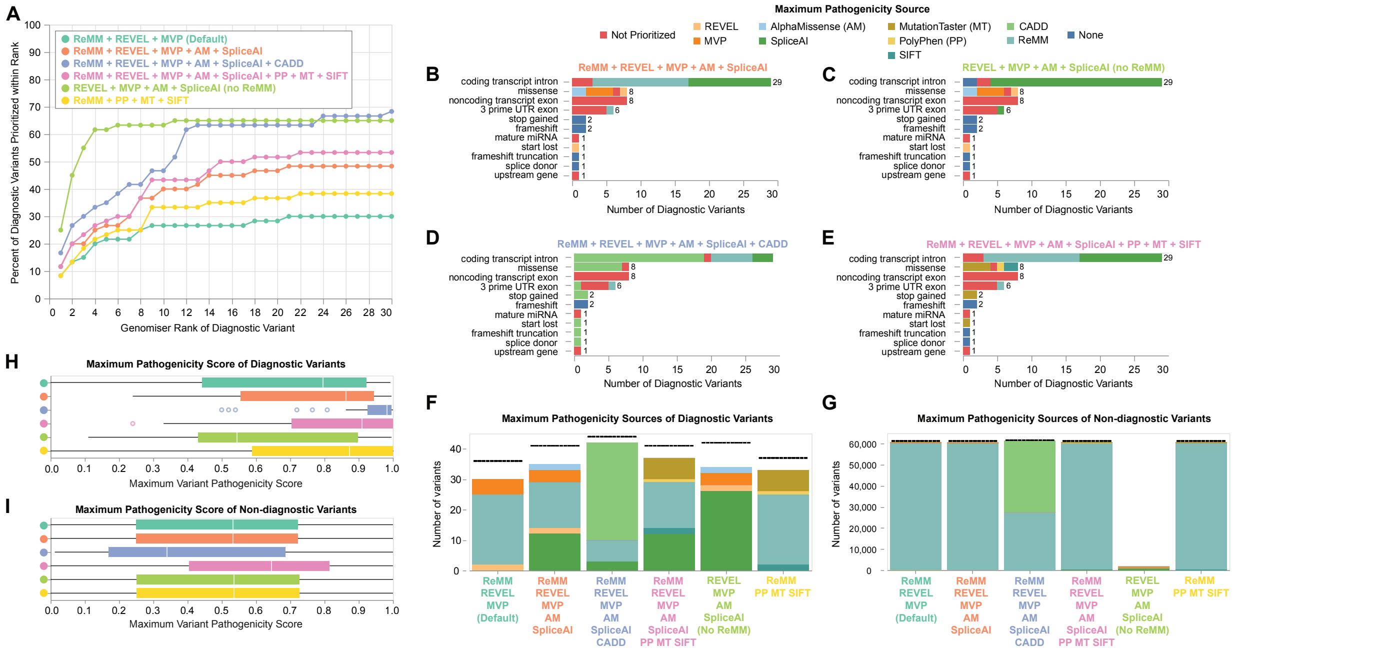


**Supplementary Figure 8: Evaluation of variant pathogenicity prediction score sources in WGS Genomiser cohort.**

All data represent Genomiser run on filtered VCFs (Methods) using hiPHIVE human-only gene:phenotype annotations.

(**A**) Average Genomiser performance across all variants under different combinations of variant pathogenicity prediction sources. X-axis indicates the rank of the diagnostic variant in the prioritized Genomiser results. Y-axis represents the percent of diagnostic variants (out of 60) that are ranked at x or lower using the selected variant pathogenicity prediction tools represented by color.

(**B-E**) Diagnostic variants' maximum pathogenicity score *source* broken down by variant class. X-axes represent the number of variants for each variant class on the y-axis (totaling to 60). Bar color represents the prediction source that provided the maximum pathogenicity score for each variant under different source combinations listed in title (top right legend). "None" represents variants that were prioritized, but do not receive a score from any selected pathogenicity score sources.

(**F-G**) Breakdown of maximum variant pathogenicity prediction score *sources* for all prioritized (**F**) diagnostic or (**G**) non-diagnostic variants under different combinations of sources (x-axis, legend in **A**). Bar color represents the prediction source that provided the maximum pathogenicity score for the number of variants represented on the y-axis (top right legend). Dashed lines represent the total number of diagnostic variants prioritized across the cohort under each run type (maximum=60 for **F**). Note that variants absent from all the selected pathogenicity sources are assigned a pathogenicity score based on their class (**Supplementary Table 2**), and therefore do not receive a maximum pathogenicity *source* (represented by this whitespace).

(**H-I**) Distribution of maximum pathogenicity *scores* of all prioritized (**H**) diagnostic or (**I**) non-diagnostic variants. Color represents the combination of variant pathogenicity score sources selected in run (legend in **A**).

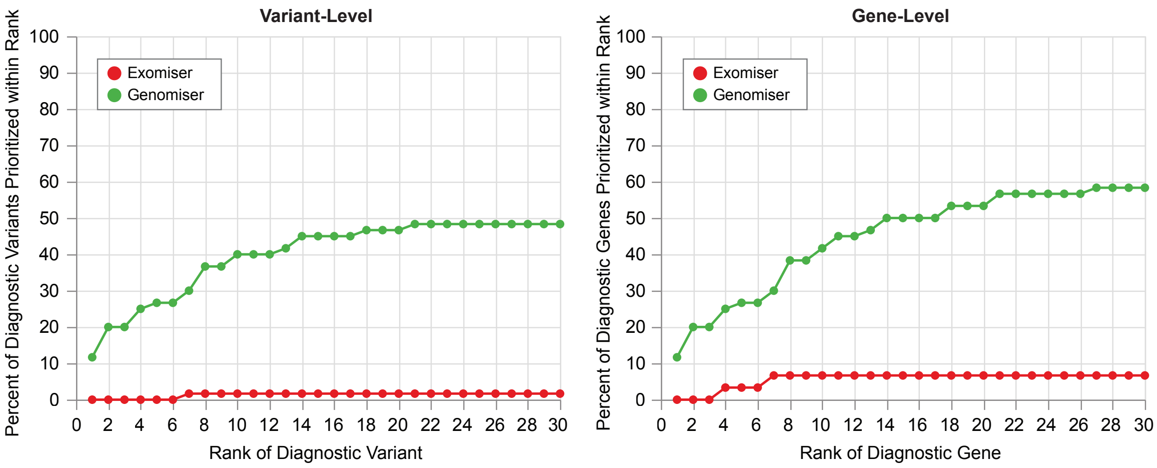


**Supplementary Figure 9**: **Exomiser performance on WGS Genomiser cohort.**
Average Exomiser (red) or Genomiser (green) performance across all variants (left) or genes (right) in WGS Genomiser cohort (n=60 variants; n=39 genes). X-axis indicates the rank of the diagnostic variant/gene in the prioritized Exomiser/Genomiser results. Y-axis represents the percent of diagnostic variants (out of 60) or genes (out of 39) that are ranked at x or lower. All lines represent Exomiser or Genomiser run on filtered VCFs (Methods) using hiPHIVE human-only gene:phenotype annotations and REVEL, MVP, AlphaMissense, SpliceAI, (and ReMM for Genomiser) variant pathogenicity prediction sources. One of 60 variants are prioritized in the top 30 candidates at the variant-level (Methods), while two of 39 genes are prioritized in the top 30 candidates at the gene-level (Methods) when running Exomiser on the WGS Genomiser cohort. This suggests that Exomiser is still unable to recognize the correct gene by identifying alternative variants at different position(s) in the true diagnostic gene.

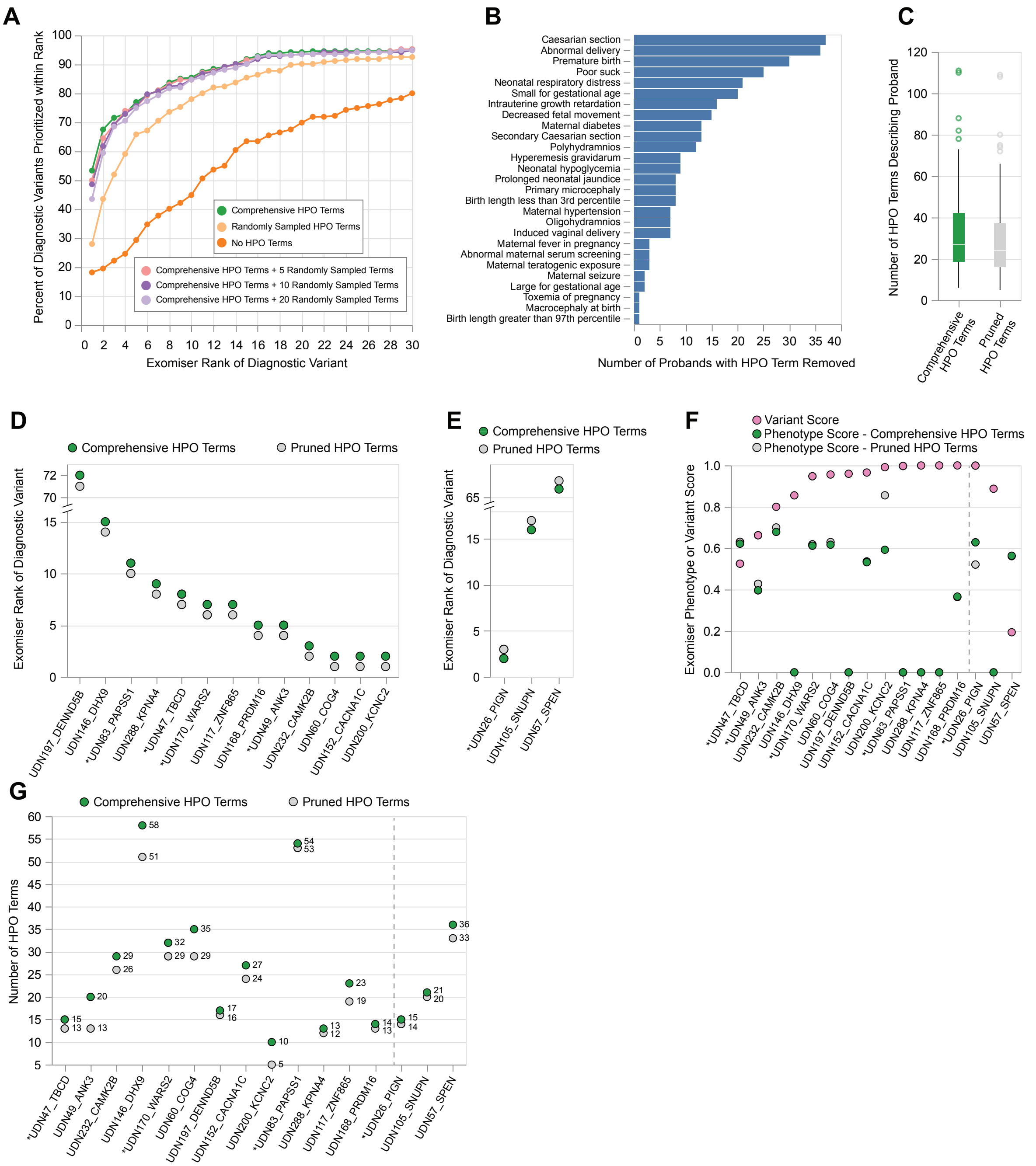


**Supplementary Figure 10: Impact of proband phenotype quality on Exomiser performance.**

(**A**) Impact of patient-specific phenotypes on Exomiser performance in WGS Exomiser cohort.

(**B**) Perinatal and prenatal HPO terms removed from the cohort of 108 WGS Exomiser pprobands to create “pruned” HPO term lists. A median of 2 terms were removed per patient (range 1-9).

Upon further investigation into these removed terms, we learned that the earliest and now obsolete phenotype collection interface for UDN cases included a separate prenatal and perinatal section with prelisted terms and checkboxes, in contrast to the clinical section where phenotype terms were added using free text matches. This illustrates the importance in HPO

term curation where biases from automated extraction or term collection interfaces may be reflected in collected HPO term sets.
(**C**) Number of HPO terms in 108 probands' comprehensive term lists in comparison to pruned lists.
Percentages in (**D**) and (**E**) calculated out of 140 variants in a cohort of 108 probands with filtered term lists.
(**D**) Running Exomiser using pruned lists as input resulted in improved ranks (by one position) for 17 (12.1%) diagnostic variants in comparison to using comprehensive term lists.
(**E**) Running Exomiser using pruned lists as input increased the rank for 4 (2.9%) diagnostic variants by one position in comparison to using comprehensive term lists.
(**F**) Exomiser phenotype scores of diagnostic variants using comprehensive or pruned HPO term lists and Exomiser variant scores for the 17 variants that improved rank (left of dashed line) and 3 variants that worsened rank (right of dashed line) following HPO term list pruning.
(**G**) Number of HPO terms in probands' comprehensive term lists in comparison to pruned lists for those whose diagnostic variants changed rank following pruning.
* denotes compound heterozygous diagnosis (2 variants in labeled gene).
All data represent Exomiser run on filtered VCF with human-only hiPHIVE phenotype prioritization and REVEL, MVP, AlphaMissense, and SpliceAI variant pathogenicity score sources.

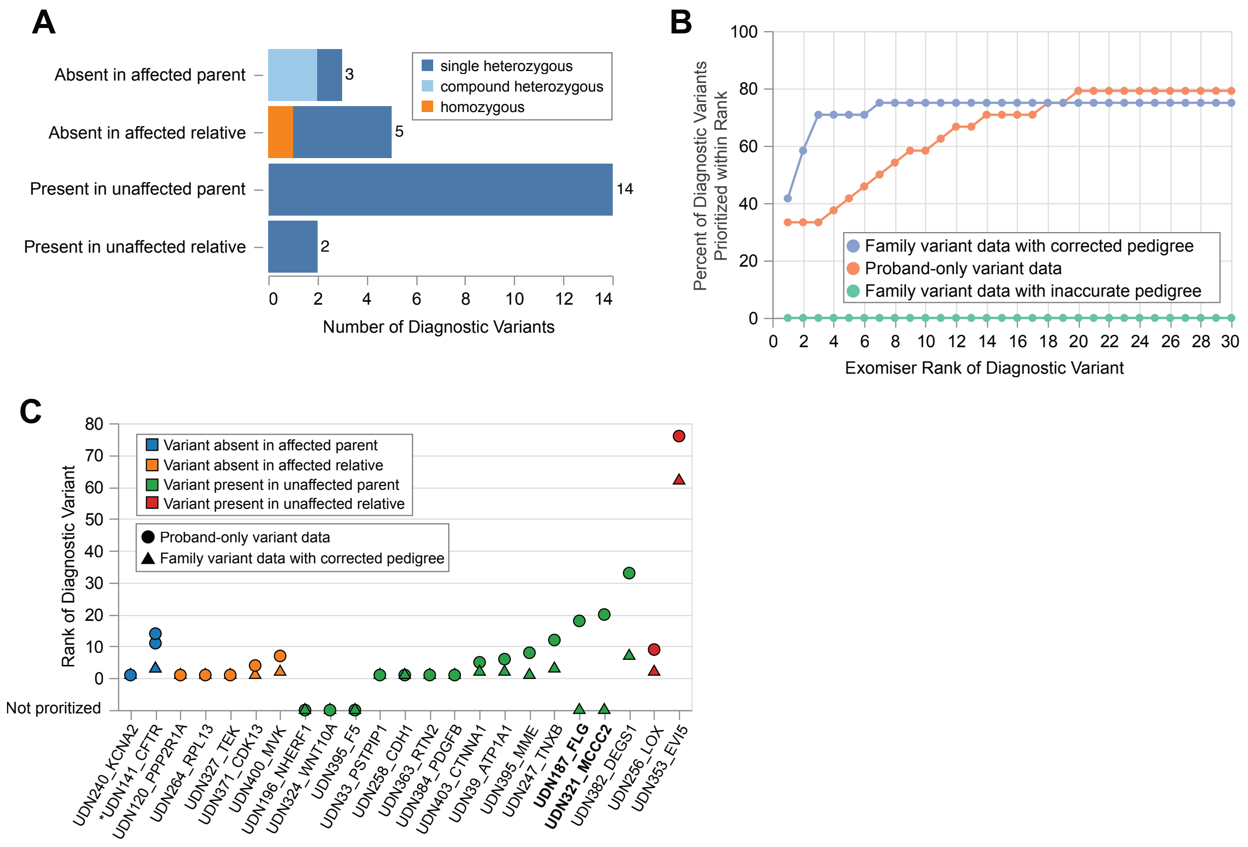


**Supplementary Figure 11: Recovering diagnostic variants through proband-only reanalysis or manual pedigree correction.**

(**A**) Breakdown of 24 diagnostic variants into four discrete categories that describe the inaccuracy in the pedigree file that led to Exomiser's failure to rank the diagnostic variant(s). These included diagnostic variants that were: (1) inherited from parents who were listed as unaffected (n=14); (2) absent in parents who were listed as affected (n=3); (3) present in non-parent relatives listed as unaffected (n=2); or (4) absent in non-parent relatives listed as affected (n=5)

(**B**) Exomiser was run on 22 families with inaccurate pedigree information that made the prioritization of the true diagnostic variant(s) impossible.

**Family variant data with corrected pedigree (blue)**: Exomiser performance using all family variant data and a manually corrected pedigree file to align the known inheritance pattern of the diagnostic variant(s).

**Proband-only variant data (orange):** Exomiser performance using only proband variant data.

**Family variant data (green)**: Exomiser performance using all family variant and pedigree data as downloaded from UDN database.

(**C**) Exomiser was run using proband-only variant data (circles) or family variant data with corrected pedigrees (triangles). Color represents the category of inaccuracy in the pedigree file as shown in (**A**). None of these 24 variants were prioritized using the complete family variant data and pedigree (not shown on graph). Bolded labels refer to the 2 variants that were recovered using proband-only data but were not prioritized when family data with corrected pedigrees were used. We note that these 2 variants would fail under our "Variant-level success with correct MOI" criteria, but pass the “Variant-level success” criteria that is our primary success measure and is most frequently reported in this manuscript (Methods).

* denotes compound heterozygous diagnosis (2 variants in labeled genes).

Exomiser was run on filtered VCFs using hiPHIVE human-only gene:phenotype annotations and REVEL, MVP, AlphaMissense, SpliceAI variant pathogenicity prediction sources with inheritance filters step enabled.

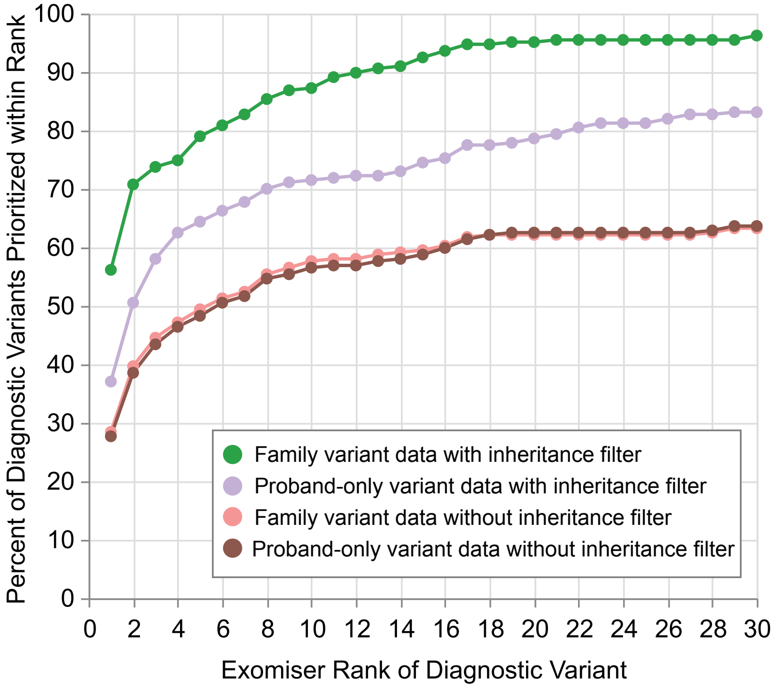


**Supplementary Figure 12: Impact of family variant data and inheritance filters on Exomiser performance.**

The WGS Exomiser cohort included 24 singleton cases (comprising 29 variants) which were excluded from this analysis. X-axis indicates the rank of the diagnostic variant in the prioritized Exomiser results. Y-axis represents the percent of diagnostic variants (out of 267) that are ranked at x or lower under the condition represented by color. Exomiser was run on filtered VCFs using hiPHIVE human-only gene:phenotype annotations and REVEL, MVP, AlphaMissense, and SpliceAI variant pathogenicity prediction sources in all conditions.

**Green** represents average Exomiser performance across all variants run on *all available family* variant data with inheritance filters step *enabled*.

**Purple** represents average Exomiser performance across all variants run on *proband-only* variant data with inheritance filters step *enabled*.

**Pink** represents average Exomiser performance across all variants run on *all available family* variant data with inheritance filters step *skipped*.

**Brown** represents average Exomiser performance across all variants run on *proband-only* variant data with inheritance filters step *skipped*. We acknowledge that this line does not exactly overlap that representing the family variant data without inheritance filter (pink) as expected. The inclusion of family data appears to slightly improve results even without the inheritance filter step, leading us to believe that Exomiser still takes some information in the provided pedigree into account even without the inheritance filtering step enabled, though, to our knowledge, this is not described in the documentation or relevant publications.

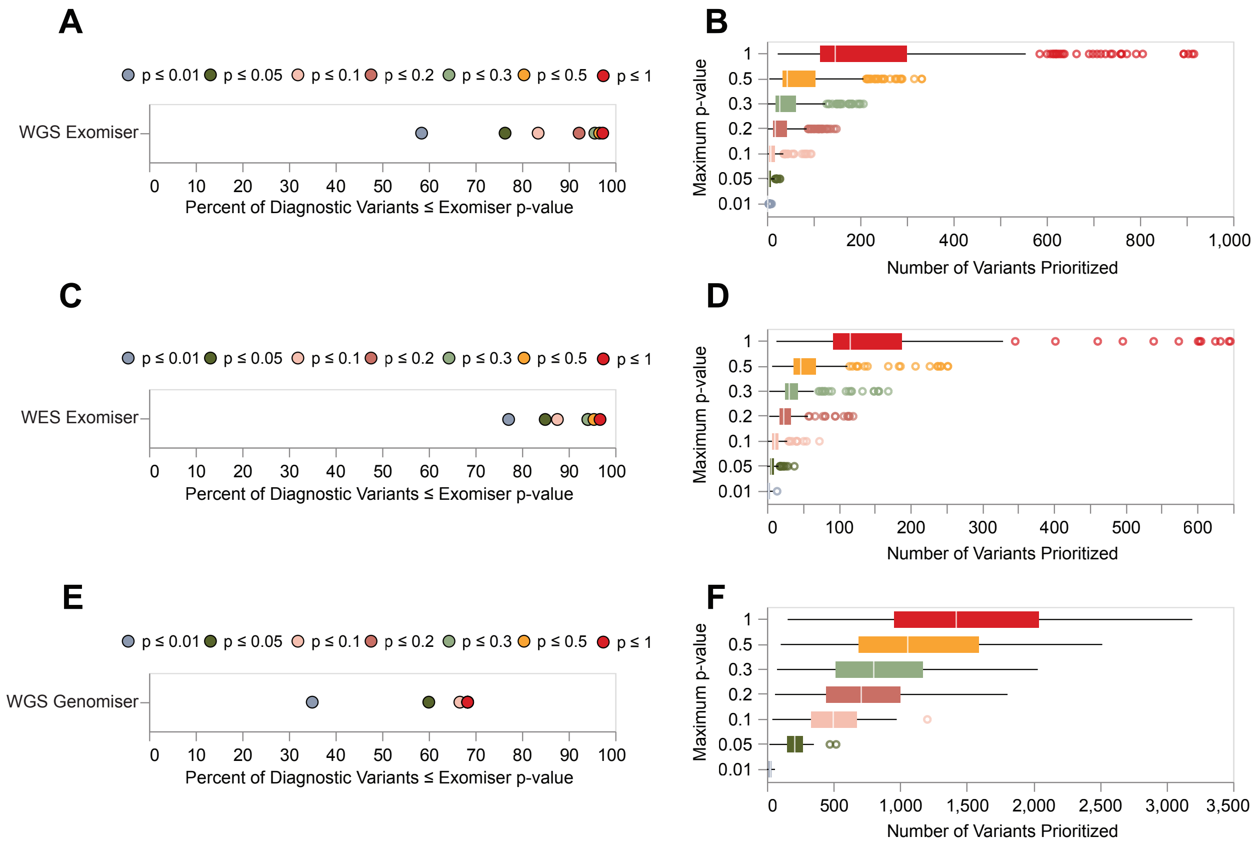


**Supplementary Figure 13: Impact of maximum p-value thresholds on length of candidate variant list and loss of diagnostic variants**.
(**A**) Percent of diagnostic variants with an Exomiser-calculated p-value ≤ threshold (color) in WGS Exomiser cohort. Maximal percentage is 97.3% (288/296 variants) (red).
(**B**) Number of variants prioritized in the Exomiser output after removing variants with p-values > threshold on y-axis (color) for 231 probands in WGS Exomiser cohort.
(**C**) Percent of diagnostic variants with an Exomiser-calculated p-value ≤ threshold (color) in WES Exomiser cohort. Maximal percentage is 96.7 (148/153 variants) (red).
(**D**) Number of variants prioritized in the Exomiser output after removing variants with p-values > threshold on y-axis (color) for 125 probands in WES Exomiser cohort.
(**E**) Percent of diagnostic variants with a Genomiser-calculated p-value ≤ threshold (color) in WGS Genomiser cohort. Maximal percentage is 68.3% (41/60 variants) (red).
(**F**) Number of variants prioritized in the Genomiser output after removing variants with p-values > threshold on y-axis (color) for 39 probands in WGS Genomiser cohort.
Exomiser was run on filtered VCFs using hiPHIVE human-only gene:phenotype annotations and REVEL, MVP, AlphaMissense, and SpliceAI variant pathogenicity prediction sources in all conditions.

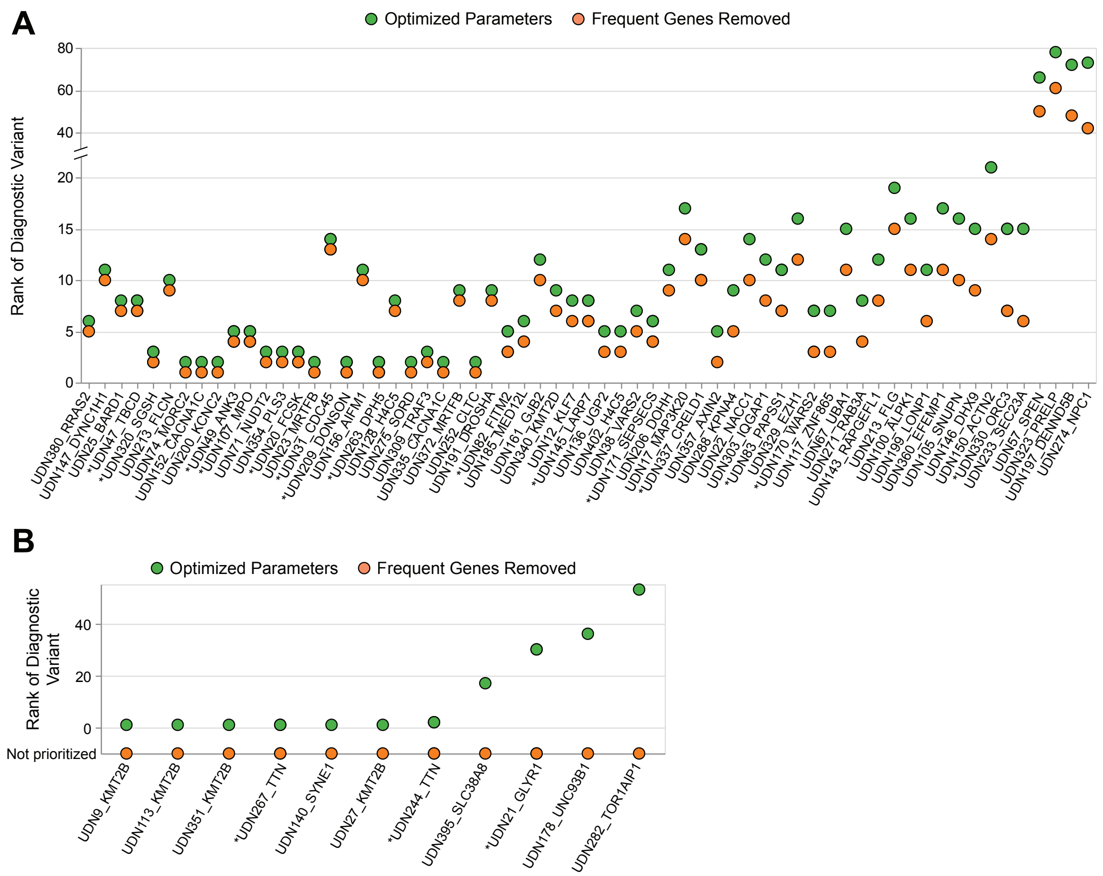


**Supplementary Figure 14: Removal of frequently ranked genes in WGS Exomiser cohort.**
(**A**) 78 variants (26.4%) improved rank after removing variants in 86 frequently prioritized genes (orange) in **Fig. 5** and subsequent reranking compared to running Exomiser using study's optimized parameters (green).
(**B**) 14 (4.7%) diagnostic variants in the benchmarking cohort were lost after removing variants in frequently prioritized genes in **Fig. 5**.
* denotes compound heterozygous diagnosis (2 variants in labeled gene).
Optimized parameters refer to running Exomiser on the filtered family VCF, hiPHIVE human-only gene:phenotype associations, and REVEL, MVP, AlphaMissense, and SpliceAI variant pathogenicity score sources.

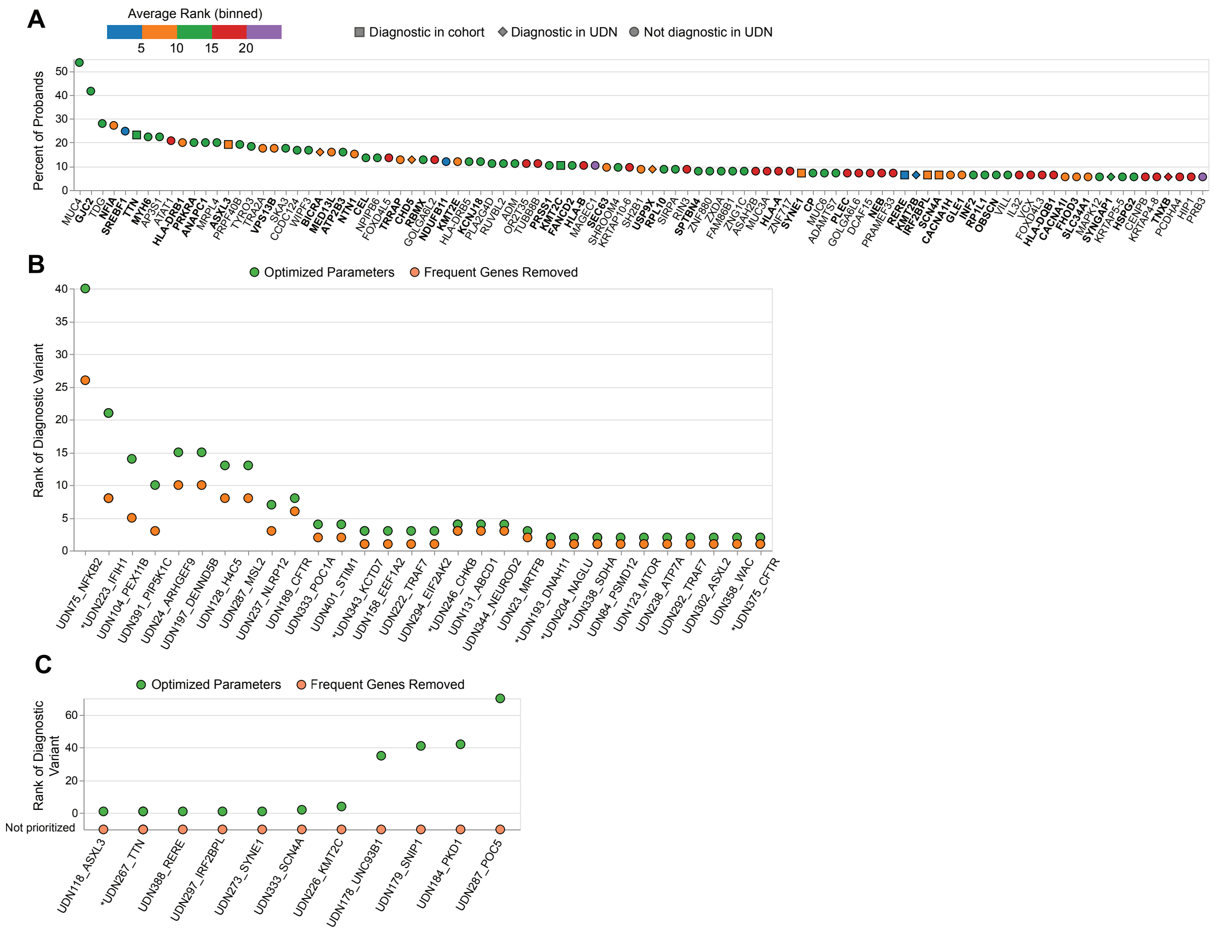


**Supplementary Figure 15: Frequently ranked genes in WES Exomiser cohort.**

(**A**) 99 genes that ranked in the top 30 candidates with $p \leq 0.3$ for at least 5% of probands in the WES Exomiser cohort. Color represents the average rank that the gene was prioritized at across all probands in the cohort. Bold font indicates OMIM genes with confirmed causal relationship to a disease (OMIM 3). Shape reflects if the gene is diagnostic in WGS Exomiser cohort (square), not diagnostic in the WGS Exomiser cohort but diagnostic in for at least one proband in the UDN consortia as a whole (diamond), or not diagnostic in the UDN consortia (circle).

(**B**) 37 variants (24.2%) improved rank after removing frequently prioritized genes (orange) in **A** compared to running Exomiser using study's optimized parameters (green).

(**C**) 12 (7.8%) diagnostic variants in the benchmarking cohort were lost after removing frequently prioritized genes in **A**.

* denotes compound heterozygous diagnosis (2 variants in labeled gene).

Optimized parameters refer to running Exomiser on the filtered family VCF (Methods), hiPHIVE human-only gene:phenotype associations, and REVEL, MVP, AlphaMissense, and SpliceAI variant pathogenicity score sources.

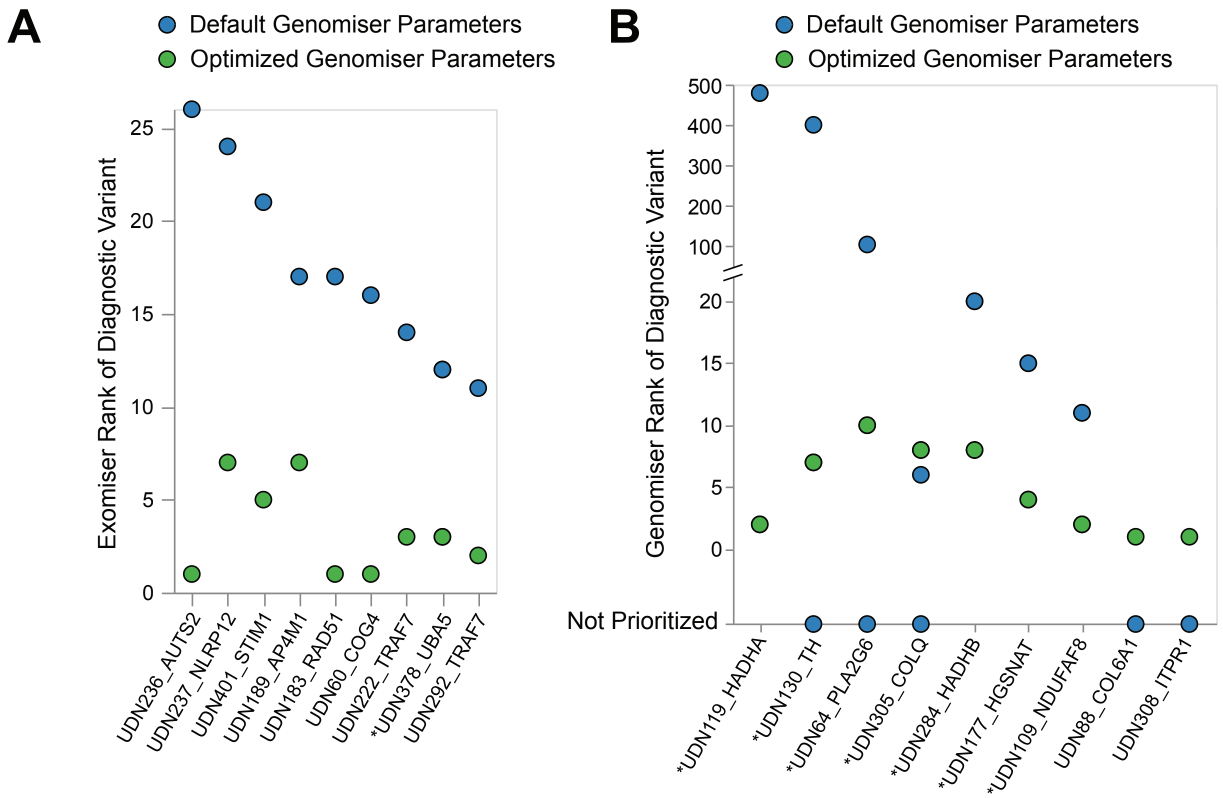


**Supplementary Figure 16: Parameter optimization shifts diagnostic variants into top ten candidates in WGS Genomiser and WES Exomiser cohorts.**

(**A**) 10 (6.5%) variants in the WES Exomiser cohort are shifted into the top ten candidates using optimized parameters (green) in comparison to default parameters (blue).

(**B**) 16 (26.7%) variants in the WGS Genomiser cohort are shifted into the top ten candidates using optimized parameters (green) in comparison to default parameters (blue).

Optimized parameters refer to running Exomiser on the *filtered* family VCF (methods), hiPHIVE human-only gene:phenotype associations, and REVEL, MVP, AlphaMissense, and SpliceAI (+ ReMM for Genomiser) variant pathogenicity score sources. Default parameters refer to running Exomiser on the filtered family VCF using hiPHIVE human, mouse, zebrafish, and PPI gene:phenotype associations and REVEL and MVP variant pathogenicity score sources.

** denotes compound heterozygous diagnosis (2 variants in labeled genes).*

### Supplementary Tables

**Supplementary Table 1: Comparison of phenotype-aware variant prioritization methods.**

| Tool | Web Access | Programmatic Access | hg38 supported | Family-based analysis | Prioritizes non-coding variants | PPI/Network Analysis Possible | Last Update (first published) |
|---|---|---|---|---|---|---|---|
| **Exomiser Genomiser** | Yes | Yes | Yes | Yes | Genomiser Only | Yes | 2024 (2014) |
| **AMELIE** | Yes | No | No | Yes | No | No | 2021 (2020) |
| **LIRICAL** | No | Yes | Yes | No | No | No | 2024 (2020) |
| **AI-MARRVEL** | Yes | Yes | Yes | Yes | Yes | ? | 2024 (2024) |
| **xRARE** | No | Yes | No | No | No | Yes | Never (2019) |
| **VARPP** | No | Yes | No | No | No | No | 2019 (2019) |
| **DeePVP** | No | Yes | No | No | No | Yes | 2018 (2019) |
| **Phenoxome** | Yes | No | No | No | No | No | Never (2019) |
| **eXtasy** | Yes | Yes | No | No | No | Yes | 2013 (2013) |
| **Phen-Gen** | No | Yes | No | Yes | Yes | Yes | Never (2014) |

Presence/absence of criteria in header. Our essential tool criteria included: (i) programmatic access to run participant VCFs in a secure environment that allows scalable analysis for hundreds of cases in parallel (as opposed to tools requiring the participant VCF(s) be uploaded to a server for manual case by case analysis); (ii) GRCh38 genome build support; (iii) multi-sample family-based analysis capability; and (iv) active maintenance with at least one update in the 3 years.

**Supplementary Table 2: Default pathogenicity scores assigned to classes of variants.**

| Variant Class | Default Score |
|---|---|
| Missense | 0.6 |
| Frameshift | 1.0 |
| Non-frameshift Indel | 0.85 |
| Nonsense | 1.0 |
| Splice Donor Acceptor | 1.0 |
| Splice Region | 0.8 |
| Synonymous | 0.10 |
| Stop Loss | 1.0 |
| Start Loss | 1.0 |
| Inversion | 0.6 |

If none of the selected variant pathogenicity prediction score sources have a prediction for a variant in the input VCF, Exomiser will automatically assign a variant pathogenicity score based on the variant's class. Values shown here were extracted from the Exomiser open-source code base.

**Supplementary Table 3: Diagnostic variants with *p*-values > 0.3 in WGS Exomiser cohort.**

| ID | UDN282 TOR1AIP1 | UDN178 UNC93B1 | UDN21 GLYR1 | UDN21 GLYR1 | UDN395 SLC38A8 |
|---|---|---|---|---|---|
| **Mode of Inheritance** | Compound heterozygous (AR) | Single heterozygous (AD) | Compound heterozygous (AR) | Compound heterozygous (AR) | Compound heterozygous (AR) |
| **Variant Class** | missense | missense | missense | missense | missense |
| **Rank** | 53 (AD) | 26 (AD) | 30 (AR) | 30 (AR) | 17 (AD) |
| **Phenotype Score – human-only** | 0.0 | 0.0 | 0.0 | 0.0 | 0.0 |
| **Variant Score** | 0.7393 | 0.6258 | 0.6078 | 0.3524 | 0.7983 |
| **Max Path Source** | AM | AM | AM | MVP | AM |
| **Combined Score** | 0.0015 | 0.0005 | 0.0001 | 0.0001 | 0.0026 |
| ***p*-value** | 0.3626 | 0.4947 | 0.6241 | 0.6241 | 0.3060 |
| **Reason for high p-value** | phenotype score 0 + low confidence AM score | phenotype score 0 + low confidence AM score | phenotype score 0 + low confidence AM score | phenotype score 0 + low MVP score | phenotype score 0 + low confidence AM score |
| **Family Structure** | Trio | Trio | Trio | Trio | Quad |
| **Number of HPO Terms** | 12 | 9 | 10 | 10 | 18 |
| **CADD score** | 24.0 | 33 | 25.1 | 19.74 | 27.0 |

AR = Autosomal recessive; AD = Autosomal dominant; AM = AlphaMissense
There were 5 (1.7%) diagnostic variants in the WGS Exomiser benchmarking cohort that had Exomiser-calculated p-values greater than 0.3. These variants were lost from the results when a *p* ≤ 0.3 threshold was applied. AM scores between 0.2-0.8 are likely less accurate, and are attributed to low confidence [34].

**Supplementary Table 4: Diagnostic variants that ranked > 30 in WGS Exomiser cohort**

| ID | UDN282 TOR1AIP1 | UDN178 UNC93B1 | UDN323 PRELP | UDN197 DENND5B | UDN274 NPC1 | UDN57 SPEN |
|---|---|---|---|---|---|---|
| Narrative (MOI) | Compound heterozygous (AR) | Single heterozygous (AD) | Single heterozygous (AD) | Single heterozygous (AD) | Single heterozygous (AD) | Single heterozygous (AD) |
| Variant Class | missense | missense | missense | missense | missense | missense |
| Rank (MOI) | 53 (AD) | 26 (AD) | 78 (AD) | 72 (AD) | 73 (AD) | 66 (AD) |
| Phenotype Score – human-only | 0.0 | 0.0 | 0.0 | 0.0 | 0.0 | 0.563 |
| Variant Score | 0.7393 | 0.6258 | 0.8087 | 0.9597 | 0.9624 | 0.1939 |
| Max Path Source | AM | AM | MVP | AM | MVP | MVP |
| Combined Score | 0.0015 | 0.0005 | 0.0028 | 0.0113 | 0.0115 | 0.0035 |
| *p*-value | 0.3626 | 0.4947 | 0.2859 | 0.1718 | 0.1436 | 0.2807 |
| Phenotype Score - all models (Rank) | 0 (88) | 0.2541 (37) | 0.5108 (37) | 0.3552 (80) | 0.2779 (89) | 0.5636 (136) |
| Reason for low rank | AR: second variant has high AF AD: Lack of gene disease associations | Lack of gene disease associations | Lack of gene disease associations | Lack of gene disease associations | Lack of gene disease associations | All variant pathogenicity predictions are low |
| Family Structure | Trio | Trio | Duo | Singleton | Singleton | Duo |
| Number of HPO Terms | 12 | 9 | 10 | 17 | 10 | 38 |
| CADD score | 24.0 | 33 | 22.5 | 22.0 | 34.0 | 15.91 |

There were seven (2.4%) diagnostic variants in the WGS Exomiser cohort that ranked beyond the top 30 candidates using our optimized parameters. Diagnostic variants beyond the top 30 candidates are unlikely to be manually reviewed by genetic counselors and clinical teams, and therefore we categorize these as “poor performance.” The poor ranking of these variants was often attributed to low predicted variant pathogenicity scores or phenotype scores, typically due to the absence of human associations linking the gene to relevant HPO terms. Although incorporating multispecies annotations (human, mouse, zebrafish, and PPI) increased the phenotype score, it did not elevate any of these variants into the top 30. These cases highlight the importance of regular updates to and the expansion of gene:phenotype association databases and variant pathogenicity predictors to improve prioritization outcomes, particularly for challenging cases. AM = AlphaMissense.

**Supplementary Table 5: Summary of diagnostic variants not prioritized by Exomiser in WGS Exomiser cohort.**

| ID | Narrative (MOI) | Variant Class | Gene Level | Variant Level; correct MOI | Variant Level | ClinVar Whitelist | Reason for Failure |
|---|---|---|---|---|---|---|---|
| UDN110 RAPSN | Compound heterozygous (AR) | missense | X | X | **X** | X | VCF Prefiltering |
| UDN110 RAPSN | Compound heterozygous (AR) | missense | X | X | **X** | X | VCF Prefiltering |
| UDN282 TOR1AIP1 | Compound heterozygous (AR) | missense | ✓ | X | **✓ (AD)** | ✓ | 2nd variant MAF > 2% |
| UDN282 TOR1AIP1 | Compound heterozygous (AR) | splice acceptor variant | ✓ | X | **X** | X | Frequency Filter MAF > 2% |
| UDN395 SLC38A8 | Compound heterozygous (AR) | missense | ✓ | X | **✓ (AD)** | ✓ | 2nd variant fails inheritance filter |
| UDN395 SLC38A8 | Compound heterozygous (AR) | missense | ✓ | X | **X** | X | Inheritance Filter |
| UDN213 FLG | Single heterozygous (AD) | stop gained | ✓ | X | **✓** | X | Only seen as AR |
| UDN295 BLK | Single heterozygous (AD) | missense | X | X | **X** | X | AD Inheritance Filter (MAF > 0.1%) |
| UDN362 CACNA1A | Single heterozygous (AD) | splice region variant | ✓ | X | **X** | X | Passed variant; non-contributing to #1 candidate gene score |
| UDN175 FLG | Single heterozygous (AD) | stop gained | X | X | **X** | ✓ | Frequency Filter MAF > 2% |
| UDN25 SERPINA1 | Homozygous (AR) | missense | ✓ | X | **X** | ✓ | Frequency Filter MAF > 2% |

Eleven variants (3.7%) in the WGS Exomiser cohort were not ranked under our most stringent success criterion, *variant-level success with correct MOI*, as described in Methods. This table summarizes each variant's outcome across all three success criteria and notes the reason for failure when applicable.

#### UDN Consortia Member List

| Full Name | Affiliation |
|---|---|
| Alyssa A. Tran | BCM Clinical |
| Arjun Tarakad | BCM Clinical |
| Ashok Balasubramanyam | BCM Clinical |
| Brendan H. Lee | BCM Clinical |
| Carlos A. Bacino | BCM Clinical |
| Daryl A. Scott | BCM Clinical |
| Elaine Seto | BCM Clinical |
| Gary D. Clark | BCM Clinical |
| Hongzheng Dai | BCM Clinical |
| Hsiao-Tuan Chao | BCM Clinical |
| Ivan Chinn | BCM Clinical |
| James P. Orengo | BCM Clinical |
| Jennifer E. Posey | BCM Clinical |
| Jill A. Rosenfeld | BCM Clinical |
| Kim Worley | BCM Clinical |
| Lindsay C. Burrage | BCM Clinical |
| Lisa T. Emrick | BCM Clinical |
| Lorraine Potocki | BCM Clinical |
| Monika Weisz Hubshman | BCM Clinical |
| Richard A. Lewis | BCM Clinical |
| Ronit Marom | BCM Clinical |
| Seema R. Lalani | BCM Clinical |
| Shamika Ketkar | BCM Clinical |
| Tiphanie P. Vogel | BCM Clinical |
| William J. Craigen | BCM Clinical |
| Jared Sninsky | BCM Clinical |
| Lauren Blieden | BCM Clinical |
| Sandesh Nagamani | BCM Clinical |
| Hugo J. Bellen | BCM MOSC |
| Michael F. Wangler | BCM MOSC |
| Oguz Kanca | BCM MOSC |

| | |
|---|---|
| Shinya Yamamoto | BCM MOSC |
| Christine M. Eng | BCM Sequencing |
| Patricia A. Ward | BCM Sequencing |
| Pengfei Liu | BCM Sequencing |
| Adeline Vanderver | CHOP |
| Cara Skraban | CHOP |
| Edward Behrens | CHOP |
| Gonench Kilich | CHOP |
| Kathleen Sullivan | CHOP |
| Kelly Hassey | CHOP |
| Ramakrishnan Rajagopalan | CHOP |
| Rebecca Ganetzky | CHOP |
| Vishnu Cuddapah | CHOP |
| Anna Raper | CHOP/UPenn |
| Daniel J. Rader | CHOP/UPenn |
| Giorgio Sirugo | CHOP/UPenn |
| Vaidehi Jobanputra | Columbia |
| Allyn McConkie-Rosell | Duke |
| Kelly Schoch | Duke |
| Mohamad Mikati | Duke |
| Nicole M. Walley | Duke |
| Rebecca C. Spillmann | Duke |
| Vandana Shashi | Duke |
| Alan H. Beggs | Harvard |
| Calum A. MacRae | Harvard |
| David A. Sweetser | Harvard |
| Deepak A. Rao | Harvard |
| Edwin K. Silverman | Harvard |
| Elizabeth L. Fieg | Harvard |
| Frances High | Harvard |
| Gerard T. Berry | Harvard |
| Ingrid A. Holm | Harvard |
| J. Carl Pallais | Harvard |
| Joan M. Stoler | Harvard |
| Joseph Loscalzo | Harvard |
| Lance H. Rodan | Harvard |
| Laurel A. Cobban | Harvard |
| Lauren C. Briere | Harvard |

| | |
|---|---|
| Matthew Coggins | Harvard |
| Melissa Walker | Harvard |
| Richard L. Maas | Harvard |
| Susan Korrick | Harvard |
| Jessica Douglas | Harvard |
| Cecilia Esteves | Harvard DMCC |
| Emily Glanton | Harvard DMCC |
| Isaac S. Kohane | Harvard DMCC |
| Kimberly LeBlanc | Harvard DMCC |
| Rachel Mahoney | Harvard DMCC |
| Shamil R. Sunyaev | Harvard DMCC |
| Shilpa N. Kobren | Harvard DMCC |
| Brett H. Graham | IU |
| Erin Conboy | IU |
| Francesco Vetrini | IU |
| Kayla M. Treat | IU |
| Khurram Liaqat | IU |
| Lili Mantcheva | IU |
| Stephanie M. Ware | IU |
| Breanna Mitchell | Mayo Clinic |
| Brendan C. Lanpher | Mayo Clinic |
| Devin Oglesbee | Mayo Clinic |
| Eric Klee | Mayo Clinic |
| Filippo Pinto e Vairo | Mayo Clinic |
| Ian R. Lanza | Mayo Clinic |
| Kahlen Darr | Mayo Clinic |
| Lindsay Mulvihill | Mayo Clinic |
| Lisa Schimmenti | Mayo Clinic |
| Queenie Tan | Mayo Clinic |
| Surendra Dasari | Mayo Clinic |
| Abdul Elkadri | MCW-CW |
| Brett Bordini | MCW-CW |
| Donald Basel | MCW-CW |
| James Verbsky | MCW-CW |
| Julie McCarrier | MCW-CW |
| Michael Muriello | MCW-CW |
| Michael Zimmermann | MCW-CW |
| Adriana Rebelo | Miami |

| | |
|---|---|
| Carson A. Smith | Miami |
| Deborah Barbouth | Miami |
| Guney Bademci | Miami |
| Joanna M. Gonzalez | Miami |
| Kumarie Latchman | Miami |
| LéShon Peart | Miami |
| Mustafa Tekin | Miami |
| Nicholas Borja | Miami |
| Stephan Zuchner | Miami |
| Stephanie Bivona | Miami |
| Willa Thorson | Miami |
| Herman Taylor | Morehouse DMCC |
| Andrea Gropman | NIH UDP |
| Barbara N. Pusey Swerdzewski | NIH UDP |
| Camilo Toro | NIH UDP |
| Colleen E. Wahl | NIH UDP |
| Donna Novacic | NIH UDP |
| Ellen F. Macnamara | NIH UDP |
| John J. Mulvihill | NIH UDP |
| Maria T. Acosta | NIH UDP |
| Precilla D'Souza | NIH UDP |
| Valerie V. Maduro | NIH UDP |
| Ben Afzali | NIH UDP, NHGRI |
| Ben Solomon | NIH UDP, NHGRI |
| Cynthia J. Tifft | NIH UDP, NHGRI |
| David R. Adams | NIH UDP, NHGRI |
| Elizabeth A. Burke | NIH UDP, NHGRI |
| Francis Rossignol | NIH UDP, NHGRI |
| Heidi Wood | NIH UDP, NHGRI |
| Jiayu Fu | NIH UDP, NHGRI |
| Joie Davis | NIH UDP, NHGRI |
| Leoyklang Petcharet | NIH UDP, NHGRI |
| Lynne A. Wolfe | NIH UDP, NHGRI |
| Margaret Delgado | NIH UDP, NHGRI |
| Marie Morimoto | NIH UDP, NHGRI |
| Marla Sabaii | NIH UDP, NHGRI |
| MayChristine V. Malicdan | NIH UDP, NHGRI |
| Neil Hanchard | NIH UDP, NHGRI |

| | |
|---|---|
| Orpa Jean-Marie | NIH UDP, NHGRI |
| Wendy Introne | NIH UDP, NHGRI |
| William A. Gahl | NIH UDP, NHGRI |
| Yan Huang | NIH UDP, NHGRI |
| Andrew Stergachis | PNW |
| Danny Miller | PNW |
| Elisabeth Rosenthal | PNW |
| Elizabeth Blue | PNW |
| Elsa Balton | PNW |
| Emily Shelkowitz | PNW |
| Eric Allenspach | PNW |
| Fuki M. Hisama | PNW |
| Gail P. Jarvik | PNW |
| Ghayda Mirzaa | PNW |
| Ian Glass | PNW |
| Kathleen A. Leppig | PNW |
| Katrina Dipple | PNW |
| Mark Wener | PNW |
| Martha Horike-Pyne | PNW |
| Michael Bamshad | PNW |
| Peter Byers | PNW |
| Runjun Kumar | PNW |
| Seth Perlman | PNW |
| Sirisak Chanprasert | PNW |
| Virginia Sybert | PNW |
| Wendy Raskind | PNW |
| Nitsuh K. Dargie | PNW |
| Chun-Hung Chan | Sanford |
| Dr. Francisco Bustos velasq | Sanford |
| Isum Ward | Sanford |
| Jason Schend | Sanford |
| Jennifer Morgan | Sanford |
| Megan Bell | Sanford |
| Miranda Leitheiser | Sanford |
| Mohamad Saifeddine | Sanford |
| Paul Berger | Sanford |
| Rachel Li | Sanford |
| Taylor Beagle | Sanford |

| | |
|---|---|
| Alexander Miller | Stanford |
| Beatriz Anguiano | Stanford |
| Beth A. Martin | Stanford |
| Brianna Tucker | Stanford |
| Chloe M. Reuter | Stanford |
| Devon Bonner | Stanford |
| Elijah Kravets | Stanford |
| Hector Rodrigo Mendez | Stanford |
| Holly K. Tabor | Stanford |
| Jacinda B. Sampson | Stanford |
| Jason Hom | Stanford |
| Jennefer N. Kohler | Stanford |
| Jennifer Schymick | Stanford |
| John E. Gorzynski | Stanford |
| Jonathan A. Bernstein | Stanford |
| Kevin S. Smith | Stanford |
| Laura Keehan | Stanford |
| Laurens Wiel | Stanford |
| Matthew T. Wheeler | Stanford |
| Meghan C. Halley | Stanford |
| Mia Levanto | Stanford |
| Page C. Goddard | Stanford |
| Paul G. Fisher | Stanford |
| Rachel A. Ungar | Stanford |
| Raquel L. Alvarez | Stanford |
| Sara Emami | Stanford |
| Shruti Marwaha | Stanford |
| Stephen B Montgomery | Stanford |
| Suha Bachir | Stanford |
| Tanner D Jensen | Stanford |
| Taylor Maurer | Stanford |
| Terra R. Coakley | Stanford |
| Euan A. Ashley | Stanford DMCC |
| Ali Al-Beshri | UAB |
| Anna Hurst | UAB |
| Brandon M Wilk | UAB |
| Bruce Korf | UAB |
| Elizabeth A Worthey | UAB |

| | |
|---|---|
| Kaitlin Callaway | UAB |
| Martin Rodriguez | UAB |
| Tammi Skelton | UAB |
| Tarun KK Mamidi | UAB |
| Andrew B. Crouse | UAB DMCC |
| Jordan Whitlock | UAB DMCC |
| Mariko Nakano-Okuno | UAB DMCC |
| Matthew Might | UAB DMCC |
| William E. Byrd | UAB DMCC |
| Albert R. La Spada | UCI/CHOC |
| Changrui Xiao | UCI/CHOC |
| Elizabeth C. Chao | UCI/CHOC |
| Eric Vilain | UCI/CHOC |
| Jose Abdenur | UCI/CHOC |
| Maija-Rikka Steenari | UCI/CHOC |
| Rebekah Barrick | UCI/CHOC |
| Sanaz Attaripour | UCI/CHOC |
| Suzanne Sandmeyer | UCI/CHOC |
| Tahseen Mozaffar | UCI/CHOC |
| Alden Huang | UCLA |
| Andres Vargas | UCLA |
| Bianca E. Russell | UCLA |
| Brent L. Fogel | UCLA |
| Esteban C. Dell'Angelica | UCLA |
| George Carvalho | UCLA |
| Julian A. Martínez-Agosto | UCLA |
| Layal F. Abi Farraj | UCLA |
| Manish J. Butte | UCLA |
| Martin G. Martin | UCLA |
| Naghmeh Dorrani | UCLA |
| Neil H. Parker | UCLA |
| Rosario I. Corona | UCLA |
| Stanley F. Nelson | UCLA |
| Yigit Karasozen | UCLA |
| Aaron Quinlan | University of Utah |
| Alistair Ward | University of Utah |
| Ashley Andrews | University of Utah |
| Corrine K. Welt | University of Utah |

| | |
|---|---|
| Dave Viskochil | University of Utah |
| Erin E. Baldwin | University of Utah |
| John Carey | University of Utah |
| Justin Alvey | University of Utah |
| Laura Pace | University of Utah |
| Lorenzo Botto | University of Utah |
| Nicola Longo | University of Utah |
| Paolo Moretti | University of Utah |
| Rebecca Overbury | University of Utah |
| Russell Butterfield | University of Utah |
| Steven Boyden | University of Utah |
| Thomas J. Nicholas | University of Utah |
| Matt Velinder | University of Utah |
| Gabor Marth | University of Utah DMCC |
| Pinar Bayrak-Toydemir | University of Utah/ARUP |
| Rong Mao | University of Utah/ARUP |
| Monte Westerfield | UO MOSC |
| Brian Corner | Vanderbilt |
| John A. Phillips III | Vanderbilt |
| Kimberly Ezell | Vanderbilt |
| Lynette Rives | Vanderbilt |
| Rizwan Hamid | Vanderbilt |
| Serena Neumann | Vanderbilt |
| Ashley McMinn | Vanderbilt |
| Joy D. Cogan | Vanderbilt |
| Thomas Cassini | Vanderbilt |
| Alex Paul | WUSTL Clinical |
| Dana Kiley | WUSTL Clinical |
| Daniel Wegner | WUSTL Clinical |
| Erin McRoy | WUSTL Clinical |
| Jennifer Wambach | WUSTL Clinical |
| Kathy Sisco | WUSTL Clinical |
| Patricia Dickson | WUSTL Clinical |
| F. Sessions Cole | WUSTL DMCC |
| Dustin Baldridge | WUSTL MOSC |
| Jimann Shin | WUSTL MOSC |

| | |
|---|---|
| Lilianna Solnica-Krezel | WUSTL MOSC |
| Stephen C. Pak | WUSTL MOSC |
| Timothy Schedl | WUSTL MOSC |
| Allen Bale | Yale |
| Carol Oladele | Yale |
| Caroline Hendry | Yale |
| Emily Wang | Yale |
| Hua Xu | Yale |
| Hui Zhang | Yale |
| Lauren Jeffries | Yale |
| María José Ortuño Romero | Yale |
| Mark Gerstein | Yale |
| Michele Spencer-Manzon | Yale |
| Monkol Lek | Yale |
| Nada Derar | Yale |
| Odelya Kaufman | Yale |
| Shrikant Mane | Yale |
| Teodoro Jerves Serrano | Yale |
| Vasilis Vasiliou | Yale |
| Winston Halstead | Yale |
| Yong-Hui Jiang | Yale |
| Bruce Gelb | Mount Sinai |
| Charlotte Cunningham-Rundles | Mount Sinai |
| Eric Gayle | Mount Sinai |
| Joanna Jen | Mount Sinai |
| Louise Bier | Mount Sinai |
| Mafalda Barbosa | Mount Sinai |
| Manisha Balwani | Mount Sinai |
| Mariya Shadrina | Mount Sinai |
| Rachel Evard | Mount Sinai |
| Saskia Shuman | Mount Sinai |
| Susan Shin | Mount Sinai |
| Ayuko Iverson | Mount Sinai |
| Kirsten Blanco | UCI/CHOC |
| Richard Chang | UCI/CHOC |
